# Glyoxal Acid-Free (GAF) histological fixative is a suitable alternative to formalin – results from an open label comparative non-inferiority study

**DOI:** 10.1101/2023.05.24.23290451

**Authors:** Ales Ryska, Anna Sapino, Stefania Landolfi, Irene Sansano Valero, Santiago Ramon y Cajal, Pedro Oliveira, Paolo Detillo, Luca Lianas, Francesca Frexia, Pier Andrea Nicolosi, Tommaso Monti, Benedetta Bussolati, Caterina Marchiò, Gianni Bussolati

## Abstract

Formalin, an aqueous solution of formaldehyde, has been the gold standard for fixation of histological samples for over a century. Despite its considerable advantages, growing evidence points to objective toxicity, particularly highlighting its carcinogenicity and mutagenic effects. In 2016, European Union proposed a ban, but a temporary permission was granted in consideration of its fundamental role in the medical-diagnostic field.

In the present study, we tested an innovative fixative, Glyoxal Acid-Free (GAF) (a glyoxal solution deprived of acids), which allows optimal tissue fixation at structural and molecular level combined with the absence of toxicity and carcinogenic activity. An open label, non-inferiority, multicentric trial was performed comparing fixation of histological specimens with GAF fixative vs standard Phosphate Buffered Formalin (PBF), evaluating the morphological preservation and the diagnostic value with four binary score questions answered by both the central pathology reviewer and local centre reviewers. The mean of total score in the GAF vs PBF fixative groups was 3.7 ± 0.5 vs 3.9 ±0.3 for the central reviewer and 3.8 ± 0.5 vs 4.0 ±0.1 for the local pathologist reviewers, respectively. In terms of median value, similar results were observed between the two fixative groups, with a median value of 4.0. Data collected indicate the non-inferiority of GAF as compared to PBF for all organ tested. The present clinical performance study, performed following the international standard for performance evaluation of *in vitro* diagnostic medical devices, highlights the capability of GAF to ensure both, structural preservation and diagnostic value of the preparations.

## Introduction

Since over a century, the gold standard for fixation of histological specimens is represented by a solution of formaldehyde in water (known as formalin) as originally proposed by Ferdinand Bloom in 1893-94 [1, 2]. This fixative has notable merits, but, on the other hand, in several aspects is its performance far from ideal. The literature contains numerous reports demonstrating formalin causing some morphological changes, loss of epitopes and artifacts in genomic sequencing [3–5]. Also, the tissue fixation is slow and, in some situations, incomplete [3–5]. On top of this, environmental authorities are increasingly concerned about the objective toxicity of formalin, which, as a volatile reagent, displays allergenic, neuro-toxic and cancerogenic properties [6–8].

Substitution of formalin as the histological fixative of choice would require adoption of a non-toxic reagent ensuring identical structural and molecular preservation of tissue as provided by formalin. Several alternative fixatives have been proposed as substitutes to formalin with the aim to meet these requirements [9]. However, none of them proved so far to be non-toxic and to match the advantages provided by formalin, in particular preservation of morphology, protection of antigenic epitopes for immunohistochemistry, maintenance of relative nucleic acids integrity for molecular-genetic testing and, at the same time, lack of over-fixation risk due to prolonged exposure to the fixative [10, 11].

Glyoxal was proposed in 1943 [12] as a fixative alternative to formalin since it is a simple dialdehyde. Glyoxal does not appear to evaporate from solution and the reported Henry’s law constant of ≤3.38 × 10^−4^Pa m^3^/mol indicates that glyoxal is essentially non-volatile in its aqueous phase [13, 14]. Therefore, it is not classified as a human carcinogen, although its use may cause some adverse reactions such as irritation of skin and eyes [15, 16]. All these data provide a clear signal that glyoxal has very low toxicity while demonstrating similar reactivity towards tissue components as formalin.

Several studies described the effects of glyoxal on tissues [9, 17–19] and a variety of fixatives based on this reagent were proposed, all of them consisting of a solution of commercial glyoxal in a water-based, acid solution. However, concerns were raised discouraging the use of this fixative as an alternative to formalin [20]. Tissues fixed in acidic glyoxal show a disturbing loss of cellular details, erythrocytes lysis and microcalcification dissolution[21]. In addition, this fixation leads to technically compromised results of fluorescence *in situ* hybridization [22, 23] as well as unsatisfactory extraction and sequencing of nucleic acids [20, 24, 25].

Based on the observations above and having observed that commercially available glyoxal is strongly acidic due to the presence of glyoxylic and glycolic acids [26], Bussolati and colleagues [27] considered that this remarkable acidity may be responsible for the observed detrimental effect on tissues. Considering this hypothesis, they developed a glyoxal solution deprived of acids using ion-exchange resins (Glyoxal Acid-Free: GAF) as a substitute of formalin for structural and molecular preservation of tissue samples [27].

In the present study, the performance of GAF fixative was tested regarding its properties as an alternative to formalin fixation. We therefore designed a trial, where, in a blinded fashion, various GAF fixed tissues were compared to formalin fixed tissues regarding quality of morphological features as well as preservation of antigens, to assess the non-inferiority of GAF compared to standard formalin fixation.

## Material and methods

### Study design

The performance evaluation study was designed as an open label, non-inferiority, multicentric trial, comparing GAF (ADDAX Biosciences Srl, Torino, Italy) as experimental fixative versus formalin as the standard fixative, on histological specimens obtained from surgical biopsies of several types of tissues most frequently analysed in routine clinical practice. The 3 centers involved in the study were: the Istituto per la Ricerca e Cura del Cancro, IRCCS of Candiolo (Torino, Italy); the Hospital Universitari Vall d’Hebron; the Vall d’Hebron Barcelona Hospital Campus (Barcelona, Spain); and the Christie NHS Foundation Trust (Manchester, United Kingdom) (Fig. 1). The study was conducted in accordance with the Declaration of Helsinki (as revised in 2013), Good Clinical Practice (GCP) principles, international standard for performance evaluation study of in vitro diagnostic (IVD) medical devices ISO 20916 (*IVD* medical devices -clinical performance studies using specimens from human subjects-good study practice) and the laws and regulations of the countries where the study took place. The protocol and all the study-related documents were approved by the local Ethical Committees (IRCCS Candiolo -Prot.269/2019 -Italy; Comité de Éticade Investigación -Pr 422/2019 – Spain; Manchester Cancer Research Centre Biobank -ref: 18/NW/009 -UK) before the beginning of the trial. All patients signed an informed consent prior to their participation in the study. The monitoring activities and data management was performed by the Contract Research Organization (C.R.O.) 1MED SA (Manno, Switzerland). The study was sponsored by ADDAX Biosciences Srl.

**Fig. 1.**
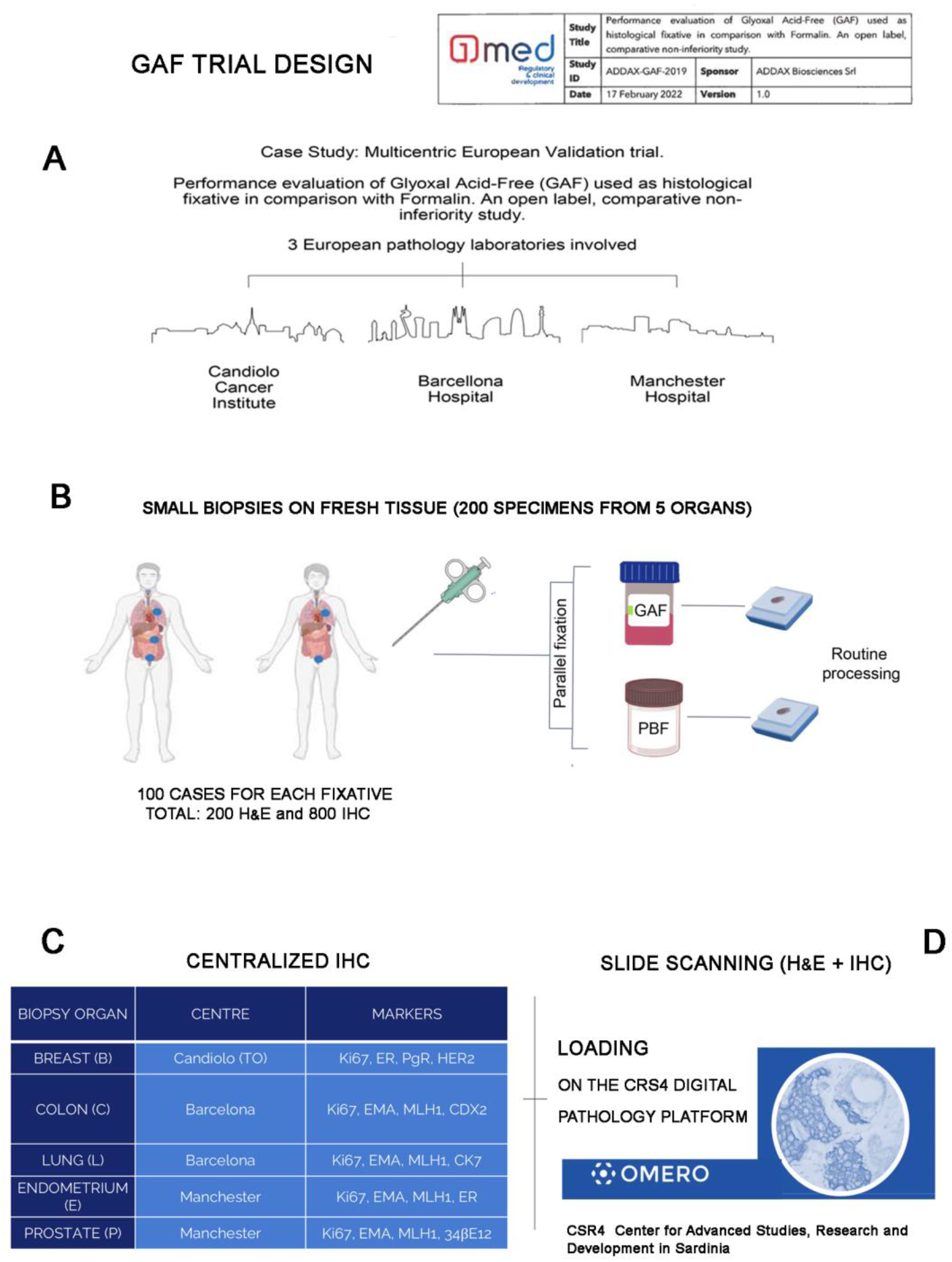
Details of the Trial development. A) The setting of the Validation Trial, as devised by the CRO (1MED). The study is multicentric, with 3 Pathology laboratories involved: the Istituto per la Ricerca e Cura del Cancro, IRCCS of Candiolo (Torino, Italy), the Hospital Universitari Vall d’Hebron; Vall d’Hebron Barcelona Hospital Campus, Barcelona (Spain) and the The Christie NHS Foundation Trust, Manchester, United Kingdom. B) Outline of the organization of the Trial, the number and site of the biopsy specimens and the type of fixation. C) Staining of tissue sections with H&E and different immune-histochemical (IHC) markers. For each of the 5 different organs, 4 IHC markers (selected among the most currently performed) were stained using a Ventana apparatus and currently adopted reagents, and processes (see IHC Table in Supplementary Material) D. Up-loading of scanned images on the CRS4 Digital Pathology Platform.

The whole and official data on the study design and conduction, statistical analysis and conclusions are presented as Supplementary data: Clinical Performance Study Report. The primary endpoint of the study was to demonstrate the non-inferiority of GAF fixation as compared to formalin fixation by comparing the total score obtained in the GAF group compared with the total score calculated in Phosphate Buffered Formalin (PBF) group on morphological preservation and diagnostic value questions answered by central pathology reviewer. The score was obtained from the answers to 4 questions. The answer to each question had a binary score (0 or 1), as detailed below. In addition, the secondary performance endpoints were the total score obtained in the GAF group compared with total score calculated in the PBF group, to be answered by local center reviewers on morphological preservation, diagnostic value, and satisfaction.

### Sample collection, fixation, and analyses

Small samples were obtained (by core needle or punch devices) from fresh surgical resection specimens, soon after surgical removal of breast, colon, and lung cancers and of hyperplastic and neoplastic lesions of prostate and endometrium. The resection specimens were sampled in a way which guaranteed that the routine diagnostic process will not be anyhow compromised. The samples (1 up to 3 mm size) were immersed at room temperature alternatively in phosphate buffered formalin (PBF, using the fixative routinely employed in the centres) and in the GAF fixative. After due time (6 h for PBF; 3 h for GAF) the specimens were transferred to alcohol, routinely processed and embedded in paraffin (Fig. 1B). Sections from the paraffin blocks were stained by haematoxylin and eosin (H&E) in the local centres, then the sections (one H&E-stained slide plus 10 unstained sections) were sent to the reference Laboratory of Pathological Anatomy and Histology of the University of Turin (Italy) for immunohistochemical (IHC) staining. The following IHC tests were performed in parallel using a Ventana BenchMark Ultra immunostainer (Ventana Medical System; Oro Valley, Arizona, USA): for breast specimens: Ki67, ER, PgR, HER2; for colon specimens: Ki67, EMA, MLH1, CDX2; for lung specimens: Ki67, EMA, MLH1, CK7; for endometrium: Ki67, EMA, MLH1, CK7; for prostate: Ki67, EMA, MLH1, 34βE12 (Fig. 1C). IHC reagents were purchased from Roche Tissue Diagnostics (Basel, Switzerland). The antigen retrieval (AR) process varied slightly between PBF and GAF-fixed samples. Reagents and times employed are outlined in Suppl. Table I. Overall, 200 specimens were collected (as 2 different samples from each case were fixed in PBF and in GAF): 90 breast; 52 colon; 16 endometrium; 22 lung; 20 prostate.

### Digital image blind presentation and scoring

Histological slides (200 H&E sections; 800 IHC stainings) were scanned with a Hamamatsu NanoZoomer S210 Digital Scanner at 40x magnification for the H&E slides and 20x for the IHCs. The obtained digital virtual slides were uploaded on the Digital Pathology Platform (DPP) developed by CRS4, based on OMERO [28] (Fig. 1D). Among the various features offered, the DPP serves as a virtual slides storage and management system and provides access to them via a Virtual Microscope (VM). To meet the requirements to perform the Trial, the DPP was extended with a new software module which allows to build multi-page questionnaires which can leverage on the integration with the VM. Extended methods for the Digital Pathology Platform generation are reported in Supplementary Methods.

In order to meet the requirements of impartiality in the reviewer’s opinion, images of sections of the same case, fixed either in formalin or in GAF, were randomly presented in the right vs. left side of the screen, so that it was not possible to predict which side contains which fixation. Each page of the questionnaire can show up to two VMs in parallel (one for PBF-fixed sample and the other for the GAF one), each of them configured to enable to switch between H&E and IHC slides (Fig. 2). The images were presented blind to the central reviewer center (University Hospital Hradec Kralove, Czech Republic), who was committed to answer to 4 questions, concerning the structural preservation of the tissue, of the nuclei, of the cytoplasm and finally on the general diagnostic value of each slide. Specifically, the questions were:

1. How do you estimate the structural preservation of the tissue?
2. How do you estimate the preservation of the nuclei?
3. How do you estimate the preservation of the cytoplasm?
4. How do you estimate the diagnostic value of these preparations?

**Fig. 2.**
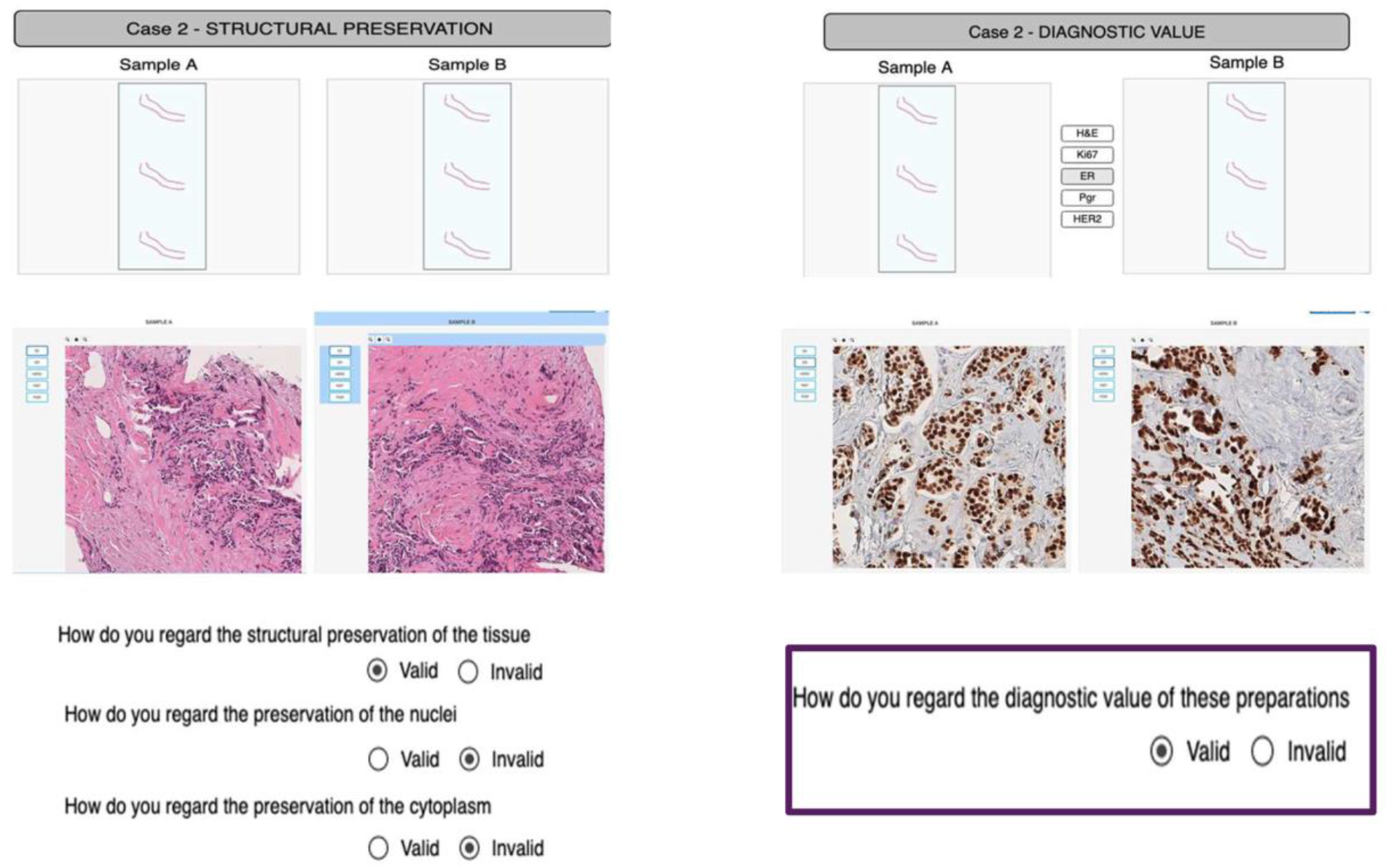
Presentation of scanned slides to the Reviewers. The program allows to visualize in parallel two images of the H&E and IHC slides of the same case, alternatively fixed in PBF and GAF. The Central Reviewer was blind to the type of fixative and answered to the questions (valid vs. invalid) on the quality and diagnostic value of the slides. The answers were directly transferred to 1MED for statistical analysis. All the of 1000 scanned images as well as the questioning posed to the Reviewers can be viewed online at: https://addax.crs4.it/datasets/1.

The answers, for statistical reasons were binary [Valid (1)/ Invalid (0)] and were directly transferred to the C.R.O. to be analyzed.

In addition, the study concerned secondary performance endpoints to be answered by local center reviewers, to test the differences between the study groups and these were concerning the score obtained from the questions: a) “Do you consider that the preparations obtained from the same case using two fixatives have the same performance?”, b) descriptive statistical analyses of IHC markers and c) reports of the pathologist’s satisfaction.

### Statistical analysis

Categorical variables were presented as numbers and percentages, and continuous variables, after evaluation of normality by applying Kolmogorov-Smirnov test, were presented as mean values, standard deviation (SD), or median value with interquartile range, as appropriate. After evaluation of the normality of the distribution of primary outcome, paired t-test or Wilcoxon test for paired data was performed to assess the difference between samples treated with GAF fixative and samples treated with PBF. If other variables were evaluated for their effect, an ANOVA model was estimated. To test the noninferiority of the GAF fixative, the null hypothesis was: H0: difference between totals. In addition, the total score calculated in the GAF group was compared with total score calculated in the PBF group on morphological preservation and diagnostic value questions answered by local center pathologists and the score obtained from “Do you consider that the preparations obtained from the same case using two fixatives have the same performance?” answered by local centers were evaluated as primary endpoint. Secondary endpoints of the study and its respective statistical considerations were the descriptive analysis of the IHC markers and results presented by the two fixative groups and the Pathologist’s satisfaction.

Sample size estimates were based on one-sided T-test assuming that the actual distribution was normal, with the assumptions of null difference between final scores calculated into each group, -2 as non-inferiority margin, 2 as standard deviation, a 10% drop-out rate, α set at 2.5% and a statistical power set at 80%. By considering the above-mentioned assumptions, a sample size of 52 biopsies per organ was estimated. Sample size estimation has been performed using SAS® proc power (SAS software version 9.4).

## Results

The details of the GAF performance Trial are described in Material and Methods and summarized in Fig. 1. Tissue samples (n. 200), alternatively fixed in PBF and in GAF, were collected from breast, colon, and lung cancers and from hyperplastic and neoplastic lesions of prostate and endometrium and embedded in paraffin. The specimens (core and punch biopsies) were of 1-2 mm in size. The recommended fixation time, at room temperature, was shorter (3 hours) for GAF fixation, while kept at 6 hours for PBF, as currently recommended for small biopsies. In the 3 centers, no technical problems were encountered in the sectioning and H&E staining, while unstained sections were sent to the central laboratory for IHC staining to eliminate any influence of interlaboratory variations.

The IHC tests to be performed were selected as the frequently performed for diagnostic purposes, on samples taken from different organs (Fig. 1). For tissue sections alternatively fixed in GAF or in PBF, the same instrument and the same reagents (all from Roche) were used. Protocols were the same for most markers, but, for some antigens, optimal results in GAF-fixed specimens were obtained when a longer AR retrieval time (up to 80’) and a longer primary antibody incubation (up to 72’) were adopted. (See Suppl. Table I). Details on the IHC Protocols for the most common markers, recommended for GAF-fixed sections are presented online (see: www.addaxbio.com→Product→IHCProtocols).

In total, 1000 slides (200 H&E slides, 800 IHC slides) have been scanned, and the related files uploaded on the CRS4 Digital Pathology Platform, to be presented blindly to the central reviewer (Fig. 1D). The Digital Pathology Platform (see Suppl. methods) allowed to visualize in parallel PBF and GAF-fixed samples of the same case, so that the reviewer could build up his opinion and answer to the questions (Fig. 2). The local reviewers could as well visualize the images and answer the questions. Scans of all H&E and IHC slides collected in this trial (1000 scanned images in total), as well as the questions posed to the reviewers, can be viewed online at: https://addax.crs4.it/datasets/1.

Both, primary and secondary endpoints data reached statistical significance. The primary endpoint of this study was concerning the evaluation of morphological preservation and diagnostic value questions answered by the blind central pathology reviewer in terms of the total score obtained in the GAF-fixed preparations compared with total score calculated in PBF-fixed ones. Mean total score in the GAF group was 3.7 ± 0.5 while in the PBF group it was 3.9 ±0.3 (Table I). However, in terms of median value, similar results were observed between fixative groups, with a median value of 4.0 (IQR: 3.5-4.0) (Table I). Applying Wilcoxon Signed-Rank test to test the non-inferiority (−2.0 of non-inferiority margin), p-value is less than 0.001, and the null hypothesis of inferiority was rejected.

**Table I.** Primary and secondary Efficacy Endpoint

| Total Score | Fixative |  |  |  |
| --- | --- | --- | --- | --- |
| Overall (N=200) | GAF | PBF | Difference between fixative<sup>a</sup> | P<sup>1</sup> |
| Central reviewer |  |  |  |  |
| N | 100 | 100 | 100 | <0.001 |
| Mean ± SD | 3.7 ± 0.5 | 3.9 ± 0.3 | -0.2 ± 0.5 | |
| Median (IQR) | 4.0 (3.5-4.0) | 4.0 (4.0-4.0) | 0.0 (0.0-0.0) |  |
| Local pathologists |  |  |  |  |
| N | 100 | 100 | 100 | <0.001 |
| Mean ± SD | 3.8 ± 0.5 | 4.0 ± 0.1 | -0.2 ± 0.5 | |
| Median (IQR) | 4.0 (3.0-4.0) | 4.0 (4.0-4.0) | 0.0 (0.0-0.0) |  |

SD: Standard Deviation; IQR: Interquartile Range; <sup>a</sup> Difference between fixative is defined as GAF total score – PBF total score; P<sup>1</sup>: p value from Wilcoxon Signed-Rank to test non-inferiority of GAF.

The non-inferiority of GAF fixative to the reference PBF was achieved for all organs tested (breast, colon, lung, endometrium, and prostate) (Fig. 3 and Supplementary data).

**Fig. 3.**
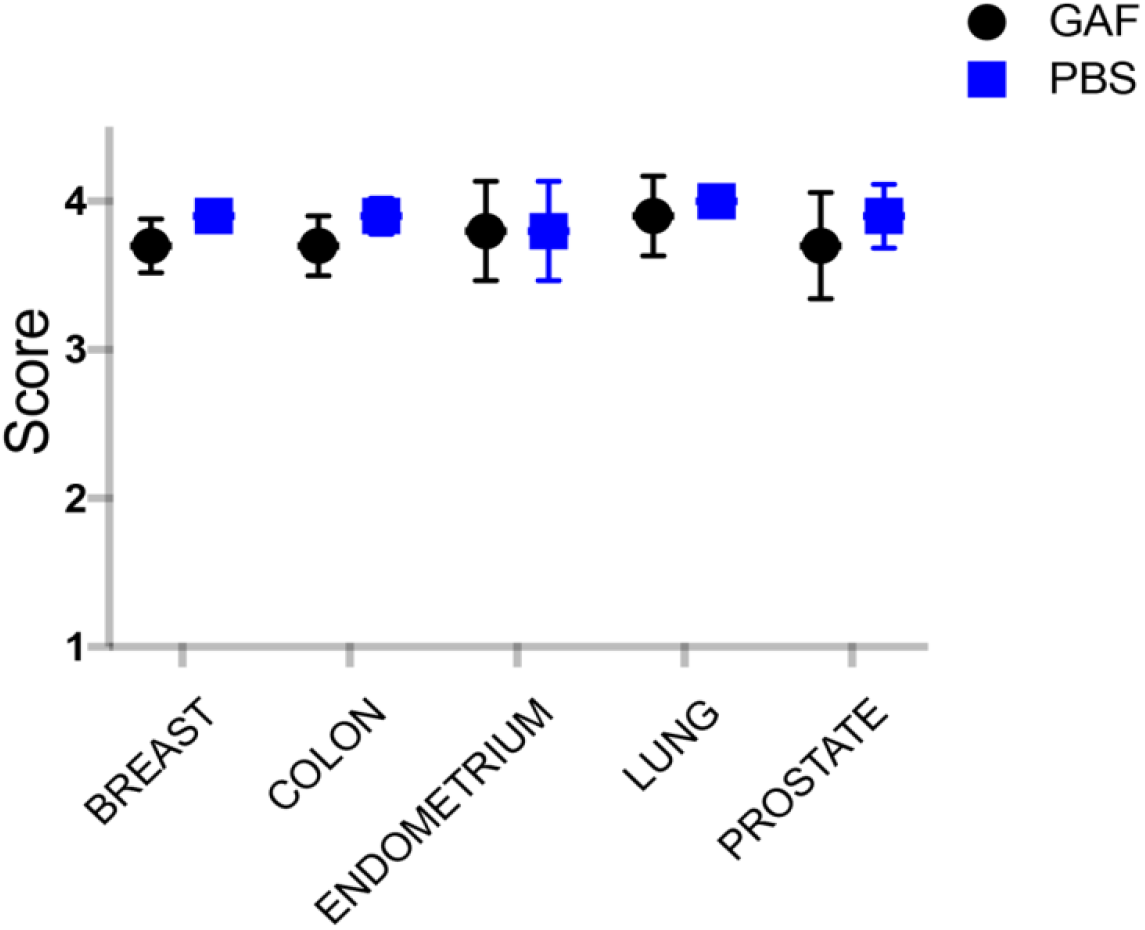
Organ-by-organ score on morphological preservation and diagnostic value (max score: 4) by the blind central reviewer. Plots show the mean ± SD scores performed on all H&E and IHC slides collected (1000 scanned images in total). N. of samples analyzed: breast: 90, colon: 52, endometrium: 16, lung: 22, prostate:20. No significant differences were observed between fixatives.

The secondary endpoint of the study concerned the evaluation of questions answered by local center pathologists on morphological preservation and diagnostic value. In general, the scores obtained by the local pathologists were very similar to those received from the central reviewer, with mean of total score in GAF fixative group of 3.8 ± 0.5 while in PBF fixative group was 4.0 ±0.1 (Table I and Supplementary data (Performance Study Report). An additional secondary endpoint of the study was the evaluation of pathologist’s satisfaction on the technical handling of the GAF-fixed samples (only asked at local centers) and the overall mean satisfaction was considered positive, with a mean value of 9.2 ± 1.1/10. Regarding the question asked to local centers “Do you consider that the slides obtained on the same case with the two fixatives have the same performance?”, 18 (18.0%) cases were answered negatively, while all the remaining 82 (82.0%) cases were answered “Yes”.

## Discussion

This performance evaluation study was focused on tissue fixation, a critical step in histopathological processing, basically aimed to the preservation of structural and chemical components as close as possible to the original status in viable tissues. Among the factors which may hamper this goal are the type of fixative, time intervals and temperature before and after immersion into the fixative fluid, the type and size of the tissue specimen, the volume of the fixative fluid and the penetration rate, which varies in different tissues [3]. A multiplicity of factors is impacting on the results, and this is bound to impose working compromises.

The routinely used fixation reagent adopted worldwide is the 10% formalin, a 4% formaldehyde solution in water, buffered to neutrality with 0.1 M Phosphate buffer, hence the term PBF. The microscopical patterns at the base of the morphological judgement of pathologists, ultimately leading to histopathological diagnoses, were formed along the years on PBF fixed tissues, building up standards for “optimal” fixation leading the educated eye to formulation of uniform and well-established diagnostic criteria. Additional factors impacting on the choice of a fixative are the speed of diffusion into the tissue, the stability, and the potential risk of over-fixation. The effect of type and time of fixation on processing, paraffin embedding, sectioning and staining are additional factors impacting on the pre-analytical process.

In recent years IHC procedures were found to be compatible with formalin fixation since antigen retrieval procedures (originally introduced by Shi et al [29]) allow restitution of most epitopes required for IHC detection. In addition, molecular biology procedures, focused on nucleic acid (DNA and RNA) analysis are as well feasible on material extracted from formalin fixed paraffin embedded tissue blocks [10,11].

Thus, the PBF fixation represents at present the gold standard in histopathology, but it is far from ideal due to multiple limitations. Formaldehyde is highly volatile, which implies the exposure of pathologists and associated personnel to the inhalation of toxic vapors. In addition to short-terms health damage (mainly asthma [30]), the personnel exposed to this reagent undergoes risks for cancer (nasopharyngeal carcinoma and myeloid leukemia [6]), brain diseases (ASL and tumors [7]) and long-term detrimental effects on cognitive health [8]. The exposure risk is well known by pathologists, who, however, tend to care for environmental protection preventing short-term damage, while accepting the subtle, but more ominous, long-term risks. In Europe, pathologists comply with the temporary permission issued by the European Parliament (UE Directive 2019/983) [31] which, in lack of a valid alternative, allows the use of formalin in this setting, but recommends limitation of exposure. A histological fixative alternative to PBF should therefore possess all properties of formaldehyde (see Table II) but toxicity and carcinogenicity.

**Table II.** Main features of PBF and GAF as histological fixatives.

| Features | PBF | GAF |
| --- | --- | --- |
| State of the art | Century old experience | Not inferior |
| Penetration | Fast | Fast |
| Fixation | Slow | Slow |
| Structural preservation | Yes | Yes |
| Tissue appearance after fixation | Hard | Soft |
| Overfixation | Absent | Absent |
| IHC | Yes | Yes |
| Nucleci acid preservation | Yes | Yes |
| Fixative quality | Volatile | Not volatile |
| Vapour toxicity | Allergenic and Neurotoxic | Absent |
| Cancerogenicity | Cancerogenic | Not cancerogenic |

Several alternatives have been proposed [9], based either on the use of acidic Glyoxal or alcoholic, coagulating reagents and considerable interest was allocated to UMFIX, a methanol-based fixative [32, 33].

The present study was focused on the demonstration that GAF, a nontoxic aldehyde fixative, is a valid alternative being not inferior to formalin in each of the characteristics which make PBF the fixative of choice for diagnostic purposes. The statistically validated results of the trial were positive, leading to the conclusion that the GAF is not inferior to PBF as a tissue fixative and is therefore ready to be used as a valid alternative for fixation of various tissues in the histopathology laboratories. Similar results in various IHC tests were obtained on sections from PBF or GAF fixed tissue blocks. However, we noticed that to obtain in GAF-fixed specimens IHC results matching those observed in the PBF-fixed ones, slightly different protocols had to be used. In particular, for most markers, more intensive AR pretreatment had to be employed. This suggests that, even though the fixation mechanism (cross-linking with methylene bonds formation) is the same for both aldehyde reagents [18], the mechanism of masking of the antigenic site is partly different. To note, the currently used antigen retrieval procedure (reagent, temperature, conditions) was originally devised for formalin fixed paraffin embedded tissues, therefore it is not surprising that the retrieval protocol must be modified for tissues with different preservation of antigens.

Secondary endpoints of the study were answers to more subjective opinions, such as evaluation of pathologist’s satisfaction when fixing tissues in GAF and on the performance of the reagent (e.g.: ease of embedding, sectioning and staining). The results were still positive, but less definite, underlying the deeply established experience of pathologists with the morphology of tissues fixed in formalin.

The study was finalized on small tissue samples, thus simulating routine biopsies performed for diagnostic purposes. Similar biopsies, obtained by core, punch or forceps devices, are routinely collected for pre-surgical evaluation of breast, lung, colon, prostate, and endometrial lesions in outpatient departments outside of pathology laboratories. In such premises, lacking hoods or other protective measures, fixation of tissue fragments in formalin may result in exposing personnel to inhalation of formaldehyde vapors. Adoption of a validated non-toxic and alternative to formalin in these settings offers the chance for a “change in safety” both, diagnostic and environmental.

This study has several limitations. One of them is the fact that it did not deal with the processing of the large surgical specimens. Different departments have different procedures how the large resection specimens are fixed. In some laboratories (mainly located in hospitals) is the material transported fresh to the laboratory, where it is immediately cut-up and the tissue blocks are subsequently fixed. Other laboratories receive the entire resection specimens already fixed by immersion in formalin and these are grossed in pathology laboratories under hoods and in relatively safe and ventilated conditions. To simulate all potential variations on fixation procedures and impact of use of an alternative fixative such as GAF would thus be not easily feasible. However, there is a clear need to extend the research to comparison of PBF and GAF fixation also in larger tissue blocks.

Another factor which was not more in depth studied in this trial is variability in duration of fixation – both shortening and prolonging the fixation time. This could test if there is any potential of GAF for further shortening of the total turnaround time of histological process as well as what is the impact of prolonged GAF fixation on potential deterioration of both tissue morphology and preservation of different molecular structures (antigens, nucleic acids) due to over-fixation.

Last limitation is the fact that we did not anyhow determine the quality of nucleic acids extracted from both types of specimens (PBF vs GAF). A dedicated study is under preparation. Moreover, the performance of GAF was recently assessed in a multicentric study performed on veterinary tissues, showing that GAF not only allows good macroscopical, histological and immunohistochemical analyses of tissue samples, but also provides better molecular analyses when compared to PBF [34]. Indeed, formalin fixed paraffin embedded tissue blocks from surgical specimens are not only analyzed histologically, but also by modern molecular procedures based on clinically validated tests, bearing a critical value in addressing prognosis and treatment in the field of precision medicine [35]. In view also of the variety of tissues and tests involved, substitution of PBF in pathology labs for the fixation of surgical specimens appears at present complex and demanding. In conclusion, the present study reports the result of an open-label multicentric Trial performed following Good Clinical Practice and the international standard for performance evaluation study of *in vitro* diagnostic medical devices (ISO 20916) [36], and confirm the non-inferiority of GAF in respect to PBF, highlight the capability of GAF to ensure the structural preservation of the: tissue, nuclei, cytoplasm and the diagnostic value of the preparations.

## Data Availability

All data produced in the present work are contained in the manuscript or in the supplementary results, or in online databases whose link is present in the manuscript.

https://addax.crs4.it/datasets/1

## Statements and Declarations

PD is an employee of Addax Biosciences srl. GB and BB are co-founder of Addax Biosciences srl.

## Acknowledgments

The study was supported by Piedmont Region, European Funds for Regional Development (POR FESR 2014-2020) trough Addax Biosciences.

## Author contributions

AR, AS, SRC, PO, PAN, BB, CM and GB contributed to the study conception and design. Material preparation, data collection and analysis were performed by AR, AS, SL, ISV, SRC, PO and PD. Data storage and slide presentation was organized by LL and FF. Statistical analysis was performed by TM. GB wrote the manuscript and all authors commented on previous versions of the manuscript. All authors read and approved the final manuscript.

## Notes

### Author Declarations

The protocol and all the study-related documents were approved by the local Ethical Committees (IRCCS Candiolo, Prot.269/2019, Italy; Comite de Eticade Investigacion, Pr 422/2019, Spain; Manchester Cancer Research Centre Biobank, ref: 18/NW/009, UK) before the beginning of the trial. All patients signed an informed consent prior to their participation in the study.

