## Supplementary for "Glyoxal Acid-Free (GAF) histological fixative is a suitable alternative to formalin – results from an open label comparative non-inferiority study"

SUPPLEMENTARY DATA

Supplementary material and methods
Supplementary references
Supplementary Table I
Supplementary data set: Clinical Performance Study Report

**Supplementary material and methods**

Digital Pathology Platform

The Digital Pathology Platform, developed by the Center for Advanced Studies, Research and Development in Sardinia CRS4 (Cagliari, Italy) is a software platform focused on the annotation of Whole Slide Images (WSI) in the context of Digital Pathology. The platform supported several international remote training initiatives [1] and collaborative research studies in the field of prostate cancer [2,3], where multiple users had to study and annotate Whole Slide Images. The system has been extended in the context of the ADDAX-GAF Trial in order to support the creation and delivery of WSI-based questionnaires.

The main software components of the platform are:

- Slides repository: based on OMERO server, it allows to handle big collections of digital slides which can be directly managed by laboratory staff;
- Virtual Microscope: called ome_seadragon, this component, developed as a plugin for OMERO.web and based on the OpenSeadragon viewer and paper.js graphical libraries, makes it possible to access and interactively annotate slides. The VM can be easily integrated in any web platform and provides a set of built-in tools to measure and annotate slides.
- Annotation Management System: this web application provides capabilities to handle structured annotations on WSI by embedding the VM. It also provides the tools to manage the delivery of the questionnaires designed for the Trial, collect the data and export them for analysis.

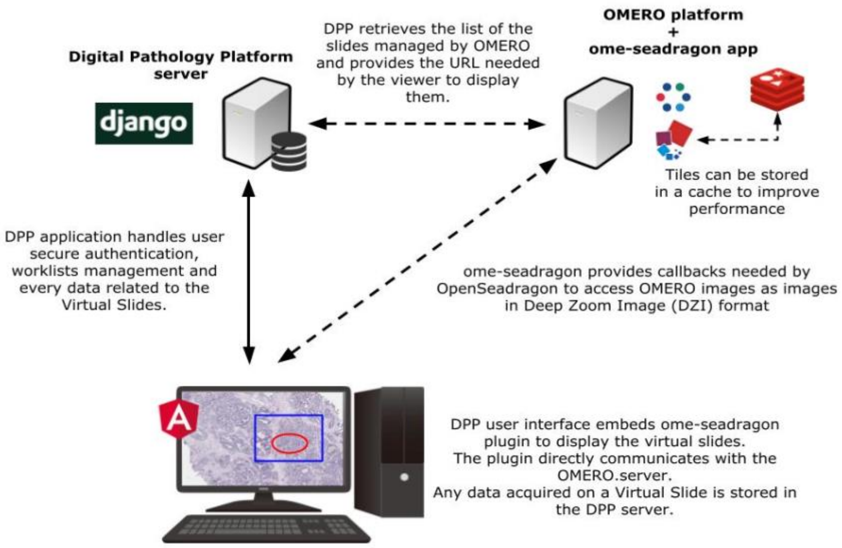

The module designed for the Trial allows to create and conduct questionnaires based on the interactive visualization of WSIs alongside a list of questions, inherent to the slides themselves, in which an arbitrary number of users can participate independently.

Each questionnaire is defined as one or more steps, each one presented as a distinct web page on the DPP; a single step can contain up to two VMs and two sets of questions. Every instance of the VM can display a single slide or a set of them in multi-page mode which allows it to easily switch to another slide of the same set without the need to create a new instance of the VM.

When two instances of the VM are displayed on the page and both of them are showing more than one slide in multi-page mode, it is possible to synchronize their views so that selecting a specific staining or Immunohistochemistry (IHC) for one sample will automatically switch to the same preparation for the other one, if available. To make this feature work, slides are aggregate as datasets in the DPP database and carefully labeled; by doing so, each IHC and staining is normalized according to a platform-defined terminology, so it is easy to compare two different sets of slides in order to check which preparations have in common and, consequently, to be able to enable synchronized exploration features.

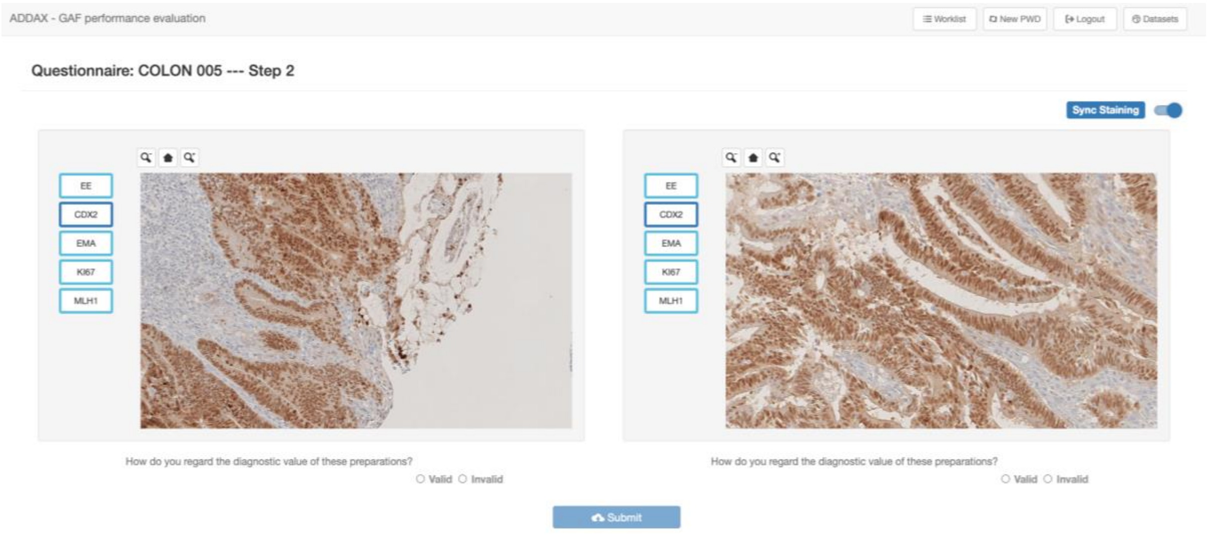

Answers to the questionnaires are collected by the DPP and made available for analysis by exporting to CSV format; to meet the Trial's requirements, each row of the file contains both central and local reviewers' responses for each case-fixative pair.

All the described software is available as Open Source under MIT license, source code is available on GitHub at the following links:

- ome_seadragon: https://github.com/crs4/ome_seadragon
- CRS4 Digital Pathology Platform: https://github.com/crs4/DigitalPathologyPlatform

Pre-configured Docker images are also available for fast and easy deployment.

**Supplementary Table I.**

| TISSUE | Fixation | Marker | A.R. | 1st Ab | Time | 2nd Ab | Time | Develop. |
|---|---|---|---|---|---|---|---|---|
| BREAST | PBF | Ki67 | CC1/52' | MiB1 | 40' | Multimer | 30' | DAB Ultraview |
| | | ER | CC1/36' | SP1 | 40' | Multimer | 30' | DAB Ultraview |
| | | PgR | CC1/36' | 1EZ | 40' | Multimer | 30' | DAB Ultraview |
| | | HER2 | CC1/36' | 435 | 40' | Multimer | 30' | DAB Ultraview |
| | GAF | Ki67 | CC1/80' | MiB1 | 60' | Multimer | 30' | DAB Optiview |
| | | ER | CC1/36' | SP1 | 60' | Multimer | 30' | DAB Ultraview |
| | | PgR | CC1/52' | 1EZ | 60' | Multimer | 30' | DAB Ultraview |
| | | HER2 | CC1/52' | 435 | 60' | Multimer | 30' | DAB Ultraview |
| COLON | PBF | Ki67 | CC1/52' | MiB1 | 40' | Multimer | 30' | DAB Ultraview |
| | | EMA | x | E29 | 10' | Multimer | 30' | DAB Ultraview |
| | | CDX2 | CC1/64' | EPR2764Y | 32' | Multimer | 30' | DAB Ultraview |
| | | MLH1 | CC1/80' | M1 | 40' | Multimer | 30' | DAB Optiview |
| | GAF | Ki67 | CC1/80' | MiB1 | 60' | Multimer | 30' | DAB Optiview |
| | | EMA | CC1/56' | E29 | 40' | Multimer | 30' | DAB Optiview |
| | | CDX2 | CC1/76' | EPR2764Y | 32' | Multimer | 30' | DAB Ultraview |
| | | MLH1 | CC1/88' | M1 | 72' | Multimer | 30' | DAB Optiview 12' ampl |
| ENDOMETRIUM | PBF | Ki67 | CC1/52' | MiB1 | 40' | Multimer | 30' | DAB Ultraview |
| | | ER | CC1/36' | SP1 | 40' | Multimer | 30' | DAB Ultraview |
| | | EMA | x | E29 | 10' | Multimer | 30' | DAB Ultraview |
| | | MLH1 | CC1/80' | M1 | 40' | Multimer | 30' | DAB Optiview |
| | GAF | Ki67 | CC1/80' | MiB1 | 60' | Multimer | 30' | DAB Optiview |
| | | ER | CC1/36' | SP1 | 60' | Multimer | 30' | DAB Ultraview |
| | | EMA | CC1/56' | E29 | 40' | Multimer | 30' | DAB Optiview |
| | | MLH1 | CC1/88' | M1 | 72' | Multimer | 30' | DAB Optiview 12' ampl |
| PROSTATE | PBF | Ki67 | CC1/52' | MiB1 | 40' | Multimer | 30' | DAB Ultraview |
| | | EMA | x | E29 | 10' | Multimer | 30' | DAB Ultraview |
| | | 34BE12 | CC1/36' | 34bE12 | 32' | Multimer | 30' | DAB Ultraview |
| | | MLH1 | CC1/80' | M1 | 40' | Multimer | 30' | DAB Optiview |
| | GAF | Ki67 | CC1/80' | MiB1 | 60' | Multimer | 30' | DAB Optiview |
| | | EMA | CC1/56' | E29 | 40' | Multimer | 30' | DAB Optiview |
| | | 34BE12 | CC1/36' | 34bE12 | 32' | Multimer | 30' | DAB Ultraview |
| | | MLH1 | CC1/88' | M1 | 72' | Multimer | 30' | DAB Optiview 12' ampl |
| LUNG | PBF | Ki67 | CC1/52' | MiB1 | 40' | Multimer | 30' | DAB Ultraview |
| | | MLH1 | CC1/80' | M1 | 40' | Multimer | 30' | DAB Optiview |
| | | EMA | x | E29 | 10' | Multimer | 30' | DAB Ultraview |
| | | CK7 | CC1/36' | SP52 | 32' | Multimer | 30' | DAB Ultraview |
| | GAF | Ki67 | CC1/80' | MiB1 | 60' | Multimer | 30' | DAB Optiview |
| | | MLH1 | CC1/80' | M1 | 40' | Multimer | 30' | DAB Optiview 12' ampl |
| | | EMA | CC1/56' | E29 | 40' | Multimer | 30' | DAB Optiview |
| | | CK7 | CC1/36' | SP52 | 32' | Multimer | 30' | DAB Ultraview |

Reagents and protocol for HIC staining. AR: antigen retrieval, amp: amplification.

| | **Study Title** | Performance evaluation of Glyoxal Acid-Free (GAF) used as histological fixative in comparison with Formalin. An open label, comparative non-inferiority study. | | |
|---|---|---|---|---|
| | **Study ID** | ADDAX-GAF-2019 | **Sponsor** | ADDAX Biosciences Srl |
| | **Date** | 17 February 2022 | **Version** | 1.0 |

### CLINICAL PERFORMANCE STUDY REPORT

#### Performance evaluation of Glyoxal Acid-Free (GAF) used as histological fixative in comparison with Formalin. An open label, comparative non-inferiority study.

| | | | |
|---|---|---|---|
| **Study dates:** | Date of start: 30 January 2020<br>Date of completion: concluded | **Report date:** | 17 February 2022 |
| **Clinical Performance Study Protocol (CPSP) identification** | ADDAX-GAF-2019 | Version: 1.0 | 19.09.2019 |
| **Investigational IVD:** | Glyoxal Acid Free Fixative (GAF). It is an innovative reagent that allows optimal tissue fixation at structural and molecular level combined with the absence of toxicity and carcinogenic activity. In Vitro diagnostic medical device.<br><br>**Indication of Use:**<br>It allows optimal tissue fixation at structural and molecular level for diagnostic purposes (e.g.: breast, prostate, colon, endometrium, and lung). | | |
| **Sponsor Name and Address:** | ADDAX Biosciences Srl,<br>Strada Mongreno 247,<br>10132 Torino, Italy<br>Phone: +39 339 6813270<br>E-Mail: | | |

*This document is confidential and is to be distributed for review only to investigators, consultants, study staff, and applicable Independent Ethics Committees National Competent Authorities or Institutional Review Boards. The contents of this document shall not be disclosed to others without written authorization from Sponsor.*

| | |
|---|---|
| **Principal investigators name, department and contact information:** | As per study design, this is a multicenter study, the centers are below reported including the Coordinating Investigator (*):<br><br>Three European Institutions are involved for the sampling:<br>Istituto per la Ricerca e Cura del Cancro (Institute for Cancer Research and Cure, IRCCS of Candiolo (Torino, Italy)). Strada Provinciale 142 km 39,5 - 10060 Candiolo (TO).<br>PI: Prof. Anna Sapino (*) (Scientific Director of the Institute, Head of the Service of Pathological Anatomy and Histology) - Tel. +39-011-9933201-3211.<br><br>Hospital Universitari Vall d'Hebron; Vall d'Hebron Barcelona Hospital Campus Passeig de la Vall d'Hebron, 119-129 - 08035 Barcelona (Spain)<br>PI: Prof. Santiago Ramon y Cajal (Head of Pathology Service) - Tel. +34 934893000 (Ext. 6934).<br><br>The Christie NHS Foundation Trust Wilmslow Road, Manchester, M20 4BX. United Kingdom.<br>PI: Dr. Pedro Soares de Oliveira Consultant in Histopathology. Dept. of Pathology. - Tel. +44-161-4463275 |
| **Coordinating center:** | Istituto per la Ricerca e cura del Cancro (Institute for Cancer Research and Cure, IRCCS of Candiolo (Torino, Italy)). Strada Provinciale 142 km 39,5 - 10060 Candiolo (TO).<br>PI: Prof. Anna Sapino (Scientific Director of the Institute, Head of the Service of Pathological Anatomy and Histology) - Tel. +39-011-9933201-3211. |
| **Author(s) of CPSR:** | 1 MED SA |
| **Compliance Statement** | The CPSP was performed in accordance with ISO 20916:2019 |

*This document is confidential and is to be distributed for review only to investigators, consultants, study staff, and applicable Independent Ethics Committees National Competent Authorities or Institutional Review Boards. The contents of this document shall not be disclosed to others without written authorization from Sponsor.*

*This document is confidential and is to be distributed for review only to investigators, consultants, study staff, and applicable Independent Ethics Committees National Competent Authorities or Institutional Review Boards. The contents of this document shall not be disclosed to others without written authorization from Sponsor.*

### 1 SUMMARY

| | |
|---|---|
| **Title:** | Performance evaluation of Glyoxal Acid-Free (GAF) used as histological fixative in comparison with Formalin. An open label, comparative non-inferiority study. |
| **Introduction:** | Glyoxal was proposed in 1943 as a fixative alternative to formalin since it is a simple di-aldehyde. As reported by Harke & Hoeffler glyoxal does not appear to evaporate from solution. Indeed, the reported Henry's law constant of ≤3.38 × 10– 4Pa m3/mol indicates that glyoxal is essentially non-volatile with regard to the aqueous phase. Glyoxal is not classifiable as a human carcinogen, nevertheless its use may cause some adverse reactions such as irritation of skin and eyes. Tumor-promoting activity of glyoxal has been reported in rats subjected to long-term exposure to this agent in drinking water. All these data are providing a clear view that glyoxal has a very low toxicity even though holding a similar reactivity to formaldehyde.<br>Several studies described the effects of glyoxal on tissues and different fixatives based on this reagent were proposed. Nevertheless, some concerns were raised discouraging the use of this fixative as an alternative to formalin. In particular, it has been claimed that glyoxal-fixed tissues show clarity of cellular details, erythrocytes are lysed and microcalcifications are dissolved. In addition, fluorescence in situ hybridization (FISH) analysis led to technically-compromised results and extraction and sequencing nucleic acids proved unsatisfactory.<br>By taking all above into consideration and having observed that commercially available glyoxal is strongly acid, Bussolati and coworkers considered that this peculiar acidity may be responsible for the observed detrimental effect on tissues. Acidification of glyoxal is likely due to its fast oxidation that leads to formation of acids, mainly glyoxilic acid, that is a very strong acid. |

*This document is confidential and is to be distributed for review only to investigators, consultants, study staff, and applicable Independent Ethics Committees National Competent Authorities or Institutional Review Boards. The contents of this document shall not be disclosed to others without written authorization from Sponsor.*

| | |
|---|---|
| | The Glyoxal Acid Free (GAF) Fixative is an innovative reagent that allows optimal tissue fixation at structural and molecular level combined with the absence of toxicity and carcinogenic activity. |
| **Purpose:** | The aim of the proposed performance evaluation study is to confirm in a large sample of histological specimens, obtained from different tissues, that an acid-free form of glyoxal (GAF) represents a novel tissue fixative by investigating morphological preservation and diagnostic value to be established on the basis of cellular details and of expression of immunohistochemical markers. |
| **Population:** | The patients enrolled will not receive any drug or intervention that could modify the clinical outcome. Patient samples will be collected, from surgical specimens of tumors arriving fresh (unfixed) from the Surgical Theatre to the Pathology labs. Samples will be obtained from pathological areas of the following organs: Breast, Colon, Uterus, Prostate and Lung. |
| **Study design** | This is an open label, non-inferiority trial, comparing GAF Fixative Vs Formalin as a fixative for histological specimens obtained from surgical biopsies, which are most frequently performed for diagnostic purposes (e.g.: breast, prostate, colon, endometrium, and lung).<br>The study is focused on the immediate preparation of the histological specimens obtained from biopsies performed on surgical samples. |
| **Number of study sites:** | 3 |
| **Statistical method used:** | Data from the study will be presented using descriptive statistics.<br>In general, categorical variables will be presented as numbers and percentages, and continuous variables, after evaluation of normality by applying Kolmogorov-Smirnov test, will be presented as mean values, standard deviation (SD), or median value with interquartile range, as appropriate.<br><br>Surgical biopsies will be included in the study according to the defined groups:<br>A. biopsies fixed in GAF;<br>B. biopsies fixed in PBF. |

*This document is confidential and is to be distributed for review only to investigators, consultants, study staff, and applicable Independent Ethics Committees National Competent Authorities or Institutional Review Boards. The contents of this document shall not be disclosed to others without written authorization from Sponsor.*

| | |
|---|---|
| | All data collected will be tabulated and represented graphically by these two fixative groups. |
| **Results of clinical performance study:** | The performance results confirmed the non-inferiority of GAF respect to PBF as highlighted by the previous interim analysis. Therefore, the GAF has the same ability of tissue fixation without toxicity and carcinogenic activity.<br>For the secondary analysis, the performance evaluations for the local laboratories was considered. Also, in this case the results confirmed the non-inferiority of GAF respect to PBF and in terms of median value similar results between fixative groups were observed.<br>The overall mean satisfaction of the local laboratories can be considered positive. |
| **Conclusions:** | Confirming the non-inferiority of GAF respect to PBF, the data of the trial highlight the capability of the investigational device to ensure the structural preservation of the: tissue, nuclei, cytoplasm and diagnostic value of the preparations (for the sections of all organs tested). These results are consistent with the rational/justification of the study and confirmed the satisfaction of local laboratories. |
| **Date of study initiation:** | 30 Jan 2020 |
| **Date of study completion:** | Site 1: 17/12/2021<br>Site 2: 16/02/2022<br>Site 3: 17/02/2022 |

#### 2 INTRODUCTION

A formaldehyde (FA) solution (Formalin) has been for many years the gold standard for fixation of histological specimens. Nevertheless, the literature contains numerous reports that FA causes some morphological changes, loss of epitopes, or mislocalization of target proteins and that it fixes the samples slowly and incompletely (1-3). On top of this, Environmental Authorities are increasingly concerned for the objective toxicity of this volatile reagent, so that a banning of formalin from 2016 has been proposed in the European Union. This has been stated by the EC Regulation n.605/2014 (4) that modifies the EC Regulation n.1272/2008 (5) defining formalin as a carcinogen (category 1B/2) and mutagen. This regulation should heavily impact on diagnostic pathology methods and procedures, even if so far the main reaction to this significant issue is limited to adoption of protective procedures, designed to prevent excessive exposure of pathology workers to formaldehyde vapours. Many other fixatives have been proposed with the aim to mitigate these problems. Among them, glutaraldehyde seems to be the most frequently used, since it fixes the samples faster and more completely than FA (6). Mixtures of FA and glutaraldehyde result in a more accurate fixation and reduce the lateral mobility of molecules (2), presumably by increasing the level of protein cross-linking. However, this fixative mixture also reduces the efficiency of immunostainings, by blocking the antibody access to epitopes, or by causing particular epitopes to unfold (7). Alcohol-based fixatives, with ice-cold methanol (2), resulted in stable fixation for a subpopulation of cellular structures (such as microtubules), but leaded to poor morphology preservation and to a loss of membranes and cytosolic proteins. Overall, the improvements in fixation induced by glutaraldehyde or methanol do not compensate for their shortcomings, thus in most cases leaving FA as the current fixative of choice. Based on the above assumptions the unmet need still to be addressed is to identify a superior alternative to FA especially since the key artifacts that were substantially negligible with the past conventional microscopy became more critical by the recent progress in super-resolution microscopy (8). In order to find a fixative that maintains high-quality immunostainings while alleviating FA problems, Bussolati G. and co-workers (9) proposed acid-free glyoxal as a substitute of formalin for structural and molecular preservation of tissue samples.

*This document is confidential and is to be distributed for review only to investigators, consultants, study staff, and applicable Independent Ethics Committees National Competent Authorities or Institutional Review Boards. The contents of this document shall not be disclosed to others without written authorization from Sponsor.*

Glyoxal was proposed in 1943 (10) as a fixative alternative to formalin since it is a simple di-aldehyde. As reported by Harke & Hoeffler (11) glyoxal does not appear to evaporate from solution. Indeed, the reported Henry's law constant of ≤3.38 × 10– 4Pa m3/mol (12) indicates that glyoxal is essentially non-volatile with regard to the aqueous phase. Glyoxal is not classifiable as a human carcinogen (13), nevertheless it use may cause some adverse reactions such as irritation of skin and eyes (13). Tumor-promoting activity of glyoxal has been reported in rats subjected to long-term exposure to this agent in drinking water (14). All these data are providing a clear view that glyoxal has a very low toxicity even though holding a similar reactivity to formaldehyde.

Several studies described the effects of glyoxal on tissues (15-17) and different fixatives based on this reagent were proposed. Nevertheless, some concerns were raised discouraging the use of this fixative as an alternative to formalin (18-19). In particular, it has been claimed that glyoxal-fixed tissues show clarity of cellular details, erythrocytes are lysed and microcalcifications are dissolved (20). In addition, fluorescence *in situ* hybridization (FISH) analysis led to technically-compromised results (21,22) and extraction and sequencing nucleic acids proved unsatisfactory (19,21, 23-25).

By taking all above into consideration and having observed that commercially available glyoxal is strongly acid, Bussolati and coworkers (9) considered that this peculiar acidity may be responsible for the observed detrimental effect on tissues. Acidification of glyoxal is likely due to its fast oxidation that leads to formation of acids, mainly glyoxilic acid, that is a very strong acid (26).

The Glyoxal Acid Free (GAF) Fixative is an innovative reagent that allows optimal tissue fixation at structural and molecular level combined with the absence of toxicity and carcinogenic activity.

Therefore, the aim of the currently proposed study is to confirm in a large sample of histological specimens, obtained from different tissues, that an acid-free form of glyoxal (GAF) represents a novel tissue fixative by investigating morphological preservation and diagnostic value to be established on the basis of cellular details and of expression of immunohistochemical marker. This is a controlled trial on the values and merits of GAF as

*This document is confidential and is to be distributed for review only to investigators, consultants, study staff, and applicable Independent Ethics Committees National Competent Authorities or Institutional Review Boards. The contents of this document shall not be disclosed to others without written authorization from Sponsor.*

a fixative compared to Formalin on the type of surgical biopsies, which are most frequently performed for diagnostic purposes (e.g.: breast, prostate, colon, endometrium, and lung).

#### 2.1 STUDY RATIONALE

In this study, the objective is to confirm, in a large sample of histological specimens obtained from different tissues, that an acid-free form of glyoxal (GAF) represents a novel tissue fixative by investigating morphological preservation and diagnostic value to be established on the basis of cellular details and of expression of immunohistochemical markers.

This is a controlled trial evaluating Glyoxal Acid-Free (GAF) compared to Phosphate buffered Formalin (PBF) as a fixative on the type of biopsies which are most frequently performed for diagnostic purposes (e.g.: breast, prostate, colon, endometrium, and lung).

**The study will evaluate the following:**

1. **Local Pathologist (each study center)**

   Will answer to the following four questions providing a score according to his/her judgment:

   1) How do you estimate the structural preservation of the tissue
   [possible answers: Valid (1)/ Invalid (0)]

   2) How do you estimate the preservation of the nuclei
   [possible answers: Valid (1)/ Invalid (0)]

   3) How do you estimate the preservation of the cytoplasm
   [possible answers: Valid (1)/ Invalid (0)]

   4) How do you estimate the diagnostic value of these preparations: (this question is focused on the H&E Preparation + the Immunohistochemical preparations).
   [possible answers: Valid (1)/ Invalid (0)]

*This document is confidential and is to be distributed for review only to investigators, consultants, study staff, and applicable Independent Ethics Committees National Competent Authorities or Institutional Review Boards. The contents of this document shall not be disclosed to others without written authorization from Sponsor.*

The answer of each question has a binary score (0 or 1). The total score of the four questions will be collected. The total score obtained by pathologists of each Center will be sent to the CRO and compared with that given by the central pathology reviewer.

In addition, the local reviewers will answer to the following questions:

Do you consider that the preparations obtained on the same case with the two fixatives have the same performance? [possible answers: Yes/No].

Were you satisfied with the use of GAF fixative during the fixation procedure? [score from 1 (not satisfied) to 10 (totally satisfied)]"

**2. Central Pathology Reviewer**

Will answer to the following four questions providing a score according to his/her judgment:
1) How do you estimate the structural preservation of the tissue
[possible answers: Valid (1)/ Invalid (0)]
2) How do you estimate the preservation of the nuclei
[possible answers: Valid (1)/ Invalid (0)]
3) How do you estimate the preservation of the cytoplasm
[possible answers: Valid (1)/ Invalid (0)]
4) How do you restimate the diagnostic value of these preparations
(this question is focused on the H&E Preparation + the Immunohistochemical preparations) [possible answers: Valid (1)/ Invalid (0)].

The answer of each question has a binary score (0 or 1). The total score of the four questions will be collected.

a) *Evaluation of the results of the relevant pre-clinical testing/assessment carried out*
Not applicable

*This document is confidential and is to be distributed for review only to investigators, consultants, study staff, and applicable Independent Ethics Committees National Competent Authorities or Institutional Review Boards. The contents of this document shall not be disclosed to others without written authorization from Sponsor.*

b) *Evaluation of clinical data that are relevant to the proposed performance evaluation study.*
Not Applicable.

#### 2.2 GUIDELINES

This study will be conducted in conformity with the ethical principles set forth by the Declaration of Helsinki, Good Clinical Practice (GCP) principles according to international standards for clinical performance studies ISO 20916:2019, the laws and regulations of the countries where the study will take place, and indemnity / insurance requirements

#### 2.3 APPROVAL OF THE INDEPENDENT ETHICS COMMITTEE OR INSTITUTIONAL REVIEW BOARD (IRB)

This investigational plan, the informed consent form (if applicable) and any other study related requested documents must be reviewed and approved by the appropriate Ethics Committee where the trial will be conducted and relevant Regulatory Authority, and any additional requirements imposed by the EC and Regulatory Authority shall be followed, if appropriate.

The clinical investigation shall not begin until the required approval/favorable opinion from the EC and Regulatory Authority have been obtained, as appropriate. Changes to the investigational plan that may increase the risk or present new risks to the subject, or that may adversely affect the validity of the trial, must be approved in writing by the Sponsor and by the Ethics Committee.

This performance evaluation study does not require any insurance policy.

### 3 DESCRIPTION OF THE IVD MEDICAL DEVICE UNDER INVESTIGATION

##### 3.1 DESCRIPTION

The Glyoxal Acid Free (GAF) Fixative is an innovative reagent that allows optimal tissue fixation at structural and molecular level combined with the absence of toxicity and carcinogenic activity. In Vitro diagnostic medical device. IVD medical device composition:

- Glyoxal Acid Free (GAF) Fixative consists of a water solution with a 2% concentration of Glyoxal (Sigma, Milan, Italy) deprived of acid by passage on ion exchange resins (Amberlyst A21, Dow Chemicals, Milan, Italy) in a Phosphate Buffer pH 7,1-7,8 and containing, as a stabilizer, Ethanol <5% , and 5% Glycol (Propylen Glycol, Chim. Strola, Turin, Italy).
- Phosphate buffered Formalin (PBF; comparator): (4% formaldehyde, in 0.1 phosphate buffer pH 7.2-7.4), of the source currently used in the reference laboratory.

##### 3.2 INTENDED USE

In Vitro diagnostic medical device with a mechanical action that is indicated:

• tissue fixation at structural and molecular level for diagnostic purposes (e.g.: breast, prostate, colon, endometrium, and lung).

##### 3.3 CHANGES TO IDV DURING THE CLINICAL INVESTIGATION

No changes to the IVD have been reported.

#### 4 CLINICAL PERFORMANCE STUDY PLAN

##### 4.1 GENERAL

**a) Description of the type of performance evaluation study to be performed with rationale for the choice**

This is a controlled trial evaluating Glyoxal Acid-Free (GAF) compared to Phosphate buffered Formalin (PBF) as a fixative on the type of biopsies which are most frequently performed for diagnostic purposes (e.g.: breast, prostate, colon, endometrium, and lung).

The objective of the proposed study is to confirm, in a large sample of histological specimens obtained from different tissues, that an acid-free form of glyoxal (GAF) represents a novel tissue fixative by investigating morphological preservation and diagnostic value to be established on the basis of cellular details and of expression of immunohistochemical markers.

**b) Description of the measures to be taken to minimize or avoid bias, including randomization and blinding/masking**

All primary and secondary outcome measures are objective, reliable, standard and verifiable.

The Trial forecasts a collection overall of 130 cases in duplicate (as 2 different biopsies from each tissue sample will be fixed in PBF and in GAF) with a forecast of 260 H&E preparations and approximately 880 IHC preparations (depending on the requests of the Pathologist in charge of the case). This means that each Centre is expected to collect about 44 cases.

After a collection of 65 cases (about 22 for each Centre) an interim analysis is forecasted.

**c) Primary and secondary endpoints, with rationale for their selection and measurement**

All primary and secondary outcome measures are objective, reliable, standard and verifiable.

<u>Primary performance Endpoint</u>

*This document is confidential and is to be distributed for review only to investigators, consultants, study staff, and applicable Independent Ethics Committees National Competent Authorities or Institutional Review Boards. The contents of this document shall not be disclosed to others without written authorization from Sponsor.*

Total score calculated in GAF group compared with total score calculated in PBF group on morphological preservation and diagnostic value questions answered by Central Pathology Reviewer

The score is obtained from the answers to 4 questions. The answer to each question has a binary score (0 or 1).

Questions:

1) How do you estimate the structural preservation of the tissue [possible answers: Valid (1)/ Invalid (0)]
2) How do you estimate the preservation of the nuclei [possible answers: Valid (1)/ Invalid (0)]
3) How do you estimate the preservation of the cytoplasm [possible answers: Valid (1)/ Invalid (0)]
4) How do you estimate the diagnostic value of these preparations (this question is focused on the H&E Preparation + the Immunohistochemical preparations) [possible answers: Valid (1)/ Invalid (0)].

Secondary performance Endpoints

The study secondary hypotheses are to test the differences between the study groups in:

- Total score calculated in GAF group compared with total score calculated in PBF group on morphological preservation and diagnostic value questions answered by local center pathologists;
- To evaluate if score obtained from "Do you consider that the preparations obtained from the same case using two fixative have the same performance?" answered by local centers can be considered as surrogate of total score obtained from 4 answers;
- Descriptive statistical analyses of IHC markers;
- Pathologist's satisfaction evaluated on local centers.

*This document is confidential and is to be distributed for review only to investigators, consultants, study staff, and applicable Independent Ethics Committees National Competent Authorities or Institutional Review Boards. The contents of this document shall not be disclosed to others without written authorization from Sponsor.*

**d) Methods and timing for assessing, recording, and analyzing variables:**

The study is focused on the immediate preparation of the histological specimens obtained from biopsies performed on surgical samples.

**e) Equipment to be used for assessing the performance evaluation study variables and arrangements for monitoring maintenance and calibration**

Not applicable.

**f) Any procedures for the replacement of subjects**

Not applicable.

#### 4.2 IVD MEDICAL DEVICE

**a) Description of the exposure to the IVD medical device:**

Fixation fluids to be used are:

**1. Phosphate buffered Formalin (PBF):**

(4% formaldehyde, in 0.1 phosphate buffer pH 7.2-7.4), of the source currently used in the reference laboratory; or

**2. Glyoxal Acid-Free Fixative (GAFF):**

The GAF fixative consists of a water solution with a 2% concentration of Glyoxal deprived of acid by passage on ion exchange resins in a Phosphate Buffer pH 7,1-7,8 and containing, as a stabilizer, Ethanol <5% , and 5% Glycol (Propylen Glycol, Chim. Strola, Turin, Italy). Phenol Red as an indicator is used to testify the basic pH of the solution.

**b) Justification of the choice of comparator**

**Glyoxal Acid-Free Fixative (GAFF)** represents a novel tissue fixative by investigating morphological preservation and diagnostic value to be established on the basis of cellular details and of expression of immunohistochemical markers.

**Phosphate buffered Formalin (PBF)** represents a golden standard.

**c) List of any other medical device or medication to be used during the Performance Evaluation Study Protocol**

Not applicable.

**d) Number of IVD medical devices to be used, together with a justification**

Only the study devices (Glyoxal Acid-Free Fixative (GAFF) or Phosphate buffered Formalin (PBF) will be used in the study, no other devices should be used.

#### 4.3 SUBJECTS

The patients enrolled will not receive any drug or intervention that could modify the clinical outcome. Patient histological specimens for this performance evaluation study will be collected in each participating centre as per local regulation (e.g. from referral biobank, from Surgical Theatre etc.).

- **Total expected duration of the performance evaluation study**

  A total duration of the study of around 6 months is foreseen from the moment of study approval until the data analysis and final results reporting.

- **Number of samples required to be included in the performance evaluation study**

  The Trial forecasts a collection overall of 130 cases in duplicate (as 2 different biopsies from each tissue sample will be fixed in PBF and in GAF) with a forecast of 260 H&E preparations and approximately 880 IHC preparations (depending on the requests of the Pathologist in charge of the case). This means that each Centre is expected to collect 44 cases, about.

  After a collection of 65 cases (about 22 for each Centre) an interim analysis is forecasted.

- **Estimated specimens' collection period**

  Expected date of start: Q4 2019;

*This document is confidential and is to be distributed for review only to investigators, consultants, study staff, and applicable Independent Ethics Committees National Competent Authorities or Institutional Review Boards. The contents of this document shall not be disclosed to others without written authorization from Sponsor.*

Expected date of completion: Q1 2020

#### 4.4 PROCEDURES

##### a) Description of all the performance evaluation study-related procedures that subjects undergo during the performance evaluation study

The patients enrolled will not receive any drug or intervention that could modify the clinical outcome. Patient histological specimens for this performance evaluation study will be collected in each participating centre as per local regulation (e.g. from referral biobank, from Surgical Theatre etc.).

The following are the procedures for the samples collection and evaluation:

The study is focused on the immediate preparation of the histological specimens obtained from biopsies performed on surgical samples.

##### 1) Collection of Samples

Samples will be collected, from fresh (unfixed) surgical specimens of tumors sent from the Surgical Theatre to the Pathology labs. Samples will be obtained from pathological areas of the following organs: Breast, Colon, Uterus, Prostate and Lung. In each of the 3 Centres, independently and in parallel, samples will be collected using core biopsy needles (gauge between 14 and 18, length 1 cm) or punch devices (size 2-4 mm).
The samples, collected in number of 1 up to 3, will therefore have a volume of 2 up to 4 $mm^3$ and will immediately be immersed in the fixation fluids.
The collected biopsies will be anonymized with an identification code, composed as follows: a code related to the Centre, a code for type of organ, a code for the samples collected in the Trial and a random code to identify the type of fixative employed (PBF or GAF).

##### 2) Processing

Fixation of the collected specimens in PBF will be processed at room temperature for 6 hours, whereas fixation of the collected specimens in GAF will be processed at room temperature for 3 hours.
Following fixation, the tissue specimens will be collected in casettes, properly labelled (see above) and immersed in Alcohol 80% for a time from 30 min. up to 48 hours to be processed for paraffin embedding using the apparatus of common use in each laboratory (either Leica or Milestone). The processing will involve passages in Alcohol 95%, followed by Absolute Alcohol, Xylene and Paraffin wax.
*NB: Specifically passages in additional fixatives, such as Formalin, have to be excluded.*

At the end of the embedding process, from the processed biopsies in paraffin blocks 4 micron thick sections will be obtained in the number of 11, using the microtome of common use in the laboratory. One section will be stained in Haematoxylin & Eosin. The other 10 unstained sections, collected in slides and properly marked with the reference number (see above), will be send to the reference Laboratory of Pathological Anatomy and Histology of the University of Turin, Italy Torino for Immunohistochemical staining. All the 11 sections (stained and unstained) will be sent to the centralized laboratory in Turin.
Once the slides have been scanned in Turin, they will be sent back to the Centre of origin, so that the Pathologist in charge of the case will answer to the same questions which will blindly be answered by the Central Reader (see below). The local Laboratory of each of the 3 Centres will retain and archive the H&E and the immunohistochemical preparations as well as the paraffin blocks.

**3) Immunohistochemical staining**

Central immunohistochemistry will be performed at the Laboratory of Pathological Anatomy and Histology of the University of Turin, Italy, in order to centralize the IHC procedures.
The unstained slides will be processed by de-paraffinization, followed by Antigen Retrieval (AR) procedures optimal for the different antigens, so that some antigens will

*This document is confidential and is to be distributed for review only to investigators, consultants, study staff, and applicable Independent Ethics Committees National Competent Authorities or Institutional Review Boards. The contents of this document shall not be disclosed to others without written authorization from Sponsor.*

not require AR, others AR at 98°C for 1 or for 3 hours, with CC1 or CC2 fluids (Ventana), alternatively with a 0.05% solution of Citraconic Anydride (Sigma) in water buffered at pH 7,4.
The sections will then be processed in Immunohistochemistry using the BenchMark ULTRA Ventana Apparatus.
Immunohistochemical reactions to be regarded as routinary in the management of biopsies of the different organs.

The following markers will be analysed:

| **Biopsy Organ** | **Centre** | **Markers (IHC Markers to be performed in every case)** |
|---|---|---|
| Breast (B) | Candiolo (TO) | Ki67, ER, PgR, HER2 |
| Colon (C) | Candiolo (TO)<br>Barcelona | Ki67, EMA, MLH1, CDX2 |
| Lung (L) | Barcelona | Ki67, EMA, MLH1, CK7 |
| Endometrium (E) | Manchester | Ki67, EMA, MLH1, ER |
| Prostate (P) | Manchester | Ki67, EMA, MLH1, 34βE12 |

**4) Scanning and Presentation**

All slides (both a slide stained with H&E and the immunohistochemical preparations of each case) will be scanned using a Hamamatsu apparatus available in the Institute in Turin. The H&E preparation of each case will be scanned at high definition (40x) while the IHC preparations will be scanned at 20x (in order not to overload the process). Each case will be scanned and presented in parallel, the PBF and the GAF fixed preparations, using the code set by the Laboratory of origin (see above). Once scanned, the images

*This document is confidential and is to be distributed for review only to investigators, consultants, study staff, and applicable Independent Ethics Committees National Competent Authorities or Institutional Review Boards. The contents of this document shall not be disclosed to others without written authorization from Sponsor.*

will be loaded on a specialized web based digital pathology platform based on OMERO (planned, produced and operated by CRS4 (Pula, CA - Italy) and then presented on a site defined as GAF Validation Trial.
Access to this site will be permitted only using: Username and Password.
Once the slides will have been scanned in Turin, they will be sent back to the Centre of origin, where they will be stored according to the standard procedures of the Center (together with the paraffin blocks).
The Pathologist in charge of the case will answer to the same questions which will blindly be answered by the Central Reader (see below).

**5) Reading**

Reading of the slides will be performed by the Central Pathology Reviewer: Prof. Ales Ryska Charles University Hradec Kralove, Czech Republic.
The Central Reviewer will not be informed of the type of fixation employed for each preparation. He will blindly open each case presented in the two variables and answer to the following 4 questions, focused solely on the H&E preparation and to a question focused on the H&E Preparation + the Immunohistochemical preparations. (see Primary & Secondary endpoints)
In addition, the PI of the three Centers will read the slides. The total score obtained by pathologists of each Center will be sent to the CRO and compared with that given by the central pathology reviewer.

**6) Sequence of events**

The 3 centres will start the collection of samples in close correlation (within short time one from the other). The first 5 cases to be collected will serve as preliminary. A meeting (either a physical meeting or a Skype conference) will be organized in order to exchange opinions and focus on possible problems encountered. At this point the collection of samples/ cases will start. An interim analysis will be performed after a collection of half of the cases.
Each of the 3 Centres is expected to collect samples of the following organs:

*This document is confidential and is to be distributed for review only to investigators, consultants, study staff, and applicable Independent Ethics Committees National Competent Authorities or Institutional Review Boards. The contents of this document shall not be disclosed to others without written authorization from Sponsor.*

the Centre in Candiolo will focus on Breast,
the Centre in Barcellona on Lung and Colon,
the Centre in Manchester on Prostate and Uterus.

**b) Description of activities performed by sponsor representatives**

No direct activity will be performed by the sponsor's representatives, except monitoring activities.

**c) Any known or foreseeable factors that may compromise the outcome of the performance evaluation study or the interpretation of results**

The anonymization of the collected biopsies will permit to avoid factors that may compromise the outcome of the performance evaluation study and the interpretation of the results.

#### 4.4 STATISTICAL CONSIDERATIONS

##### a) Analytical procedures

Data from the study will be presented using descriptive statistics. In general, categorical variables will be presented as numbers and percentages, and continuous variables, after evaluation of normality by applying Kolmogorov-Smirnov test, will be presented as mean values, standard deviation (SD), or median value with interquartile range, as appropriate.

Surgical biopsies will be included in the study according to the defined groups:

A. biopsies fixed in GAF;

B. biopsies fixed in PBF.

All data collected will be tabulated and represented graphically by these two fixative groups.

*This document is confidential and is to be distributed for review only to investigators, consultants, study staff, and applicable Independent Ethics Committees National Competent Authorities or Institutional Review Boards. The contents of this document shall not be disclosed to others without written authorization from Sponsor.*

**b) Sample size**

Because of primary endpoint of the study is based on the answer of 4 questions formulated ad hoc for the study, no bibliographical references will be considered to calculate the sample size.

For this reason, descriptive statistic of theorical distribution of difference between two final scores obtained from two groups (i.e. interquartile range) will be considered as expected effect in terms of mean difference in final score and his standard deviation.

Sample size estimates are based on one-sided T-test assuming that the actual distribution is normal.

The following assumptions have been made in order to estimate the sample size.

1. Null difference between final scores calculated into each group is considered to demonstrate the non-inferiority of GAF respects to PBF.

2. The 25th percentile of theorical distribution of differences between two final scores is equal to -2 and it is considered as non-inferiority margin.

3. The standard deviation has been calculated as suggested in Conroy R.'s guide. The highest value is maximum of the distribution (it is equal to 4) and the lowest value is the minimum value (it is equal to -4), then the standard deviation has been set on 2 points (27).

4. The significance level (α), i.e. the probability of the study detecting a false positive finding, has been set at 2.5%.

5. The statistical power, i.e. the probability of detecting an effect when the effect really exists, has been set at 80%.

By considering the above-mentioned assumptions, a sample size of 46 biopsies has been estimated.

Planning to randomize a total of 52 biopsies (26 in each fixative group) would allow for a 10% drop-out rate. Sample size estimated above will be considered just for one

*This document is confidential and is to be distributed for review only to investigators, consultants, study staff, and applicable Independent Ethics Committees National Competent Authorities or Institutional Review Boards. The contents of this document shall not be disclosed to others without written authorization from Sponsor.*

organ evaluated, because of surgical biopsies will be collected from 5 different organs (breast, colon, prostate, lung, and endometrium). Then the final sample size will be 260 surgical biopsies (130 in each group). Sample size estimation has been performed using SAS® proc power (SAS software version 9.4 (28)).

**c) Study population**

All analyses will be performed on all randomized surgical biopsies. In case of some sample will not be analyzable, this will be considered a drop-out and it will be excluded from the analysis.

**d) Performance analysis**

Primary endpoint

Primary endpoint of the study will be the total score obtained in GAF group compared with total score calculated in PBF group on morphological preservation and diagnostic value questions answered by Central Pathology Reviewer. The score is obtained from the answers to 4 questions. The answer to each question has a binary score (0 or 1).

After evaluation the normality of the distribution of primary outcome, paired t-test or Wilcoxon test for paired data will be performed to assess the difference between samples treated with GAF fixative and samples treated with PBF. If other variables will be evaluated for their effect, an ANOVA model will be estimated. To test non-inferiority of the GAF fixative, the null hypothesis will be: H0: difference between total scores from two groups of fixatives is < -2; if it will be rejected, non-inferiority of the novel fixative will be accepted.

Secondary endpoints

Secondary endpoints of the study and its respective statistical considerations will be:

1. Total score calculated in GAF group compared with total score calculated in PBF group on morphological preservation and diagnostic value questions answered by local center pathologists. This endpoint will be evaluated as primary endpoint;

*This document is confidential and is to be distributed for review only to investigators, consultants, study staff, and applicable Independent Ethics Committees National Competent Authorities or Institutional Review Boards. The contents of this document shall not be disclosed to others without written authorization from Sponsor.*

2. To evaluate if score obtained from "Do you consider that the preparations obtained from the same case using two fixative have the same performance?" answered by local centers can be considered as surrogate of total score obtained from 4 answers. This endpoint will be evaluated as primary endpoint;

3. IHC markers will be analyzed descriptively and results will be presented by two fixative groups;

4. Pathologist's satisfaction evaluated on local centers will be analyzed descriptively and results will be presented by two fixative groups.

**e) Safety analysis**

Not applicable

**f) Interim analysis**

After a collection of 65 cases (about 22 for each Centre) an interim analysis is forecasted. At this time point the study will be temporarily put on hold until the interim analysis will be performed and the scientific advisory Board will release its positive evaluation to continue. At that time the study will be restarted up to completion of all the samples required by the protocol.

**g) Procedures for reporting any deviation(s) from the original statistical plan**

A fully specified Statistical Analysis Plan (SAP) for Clinical Study Report (CSR) will be prepared before the data base lock. The contents of the SAP will include a full and detailed descriptions of the statistical methods for data analysis, as well as a detailed description of the contents of tables, listings and figures. The plan may be reviewed and updated before the start of the statistical analysis, which will start only at the end of data management activities.

**h) Specification of subgroups for analysis**

Not applicable in this study.

**i) Treatment of missing, unused or spurious data, including drop-outs and**

*This document is confidential and is to be distributed for review only to investigators, consultants, study staff, and applicable Independent Ethics Committees National Competent Authorities or Institutional Review Boards. The contents of this document shall not be disclosed to others without written authorization from Sponsor.*

**withdrawals**

Not applicable in this study.

#### 4.5 RISKS AND BENEFITS OF THE IVD MEDICAL DEVICE AND CLINICAL INVESTIGATION

*a) Anticipated clinical benefits:*

No clinical benefits are expected as the aim of the currently proposed study is to confirm in a large sample of histological specimens, obtained from different tissues, that an acid-free form of glyoxal (GAF) represents a novel tissue fixative by investigating morphological preservation and diagnostic value to be established on the basis of cellular details and of expression of immunohistochemical marker

*b) Anticipated adverse device effects:*

The adverse events listed below may occur:

- Eye contact causes irritation. Symptoms may include: redness, edema, pain and tearing.

- Skin contact may cause moderate irritation. Product contact with the skin causes sensitization (contact dermatitis). The dermatitis originates following an inflammation of the skin, which begins in the skin areas that come into repeated contact with the sensitising agent. The skin lesions may include erythema, edema, papules, vesicles, pustules, scales, fissures and exudative phenomena, which vary according to the stages of the disease and the areas affected. In the acute phase the following prevails: erythema, edema and exudation. In chronic phases the following prevails: scales, dryness, fissuration and thickening of the skin.

- Ingestion may cause health problems, which include pain with burning, nauseous and vomiting.

*c) Residual risks associated with the IVD medical device, as identified in the risk analysis*

*This document is confidential and is to be distributed for review only to investigators, consultants, study staff, and applicable Independent Ethics Committees National Competent Authorities or Institutional Review Boards. The contents of this document shall not be disclosed to others without written authorization from Sponsor.*

*report:*

Based on the risk-analysis assessment, it is considered that the device produced and used in compliance with the operating procedures described by ADDAX Biosciences srl, does not present any relevant risks (that are deemed not acceptable) for the study.

*d) Risks associated with participation in the performance evaluation study*

No risks are expected as the aim of the currently proposed study is to confirm in a large sample of histological specimens, obtained from different tissues, that an acid-free form of glyoxal (GAF) represents a novel tissue fixative by investigating morphological preservation and diagnostic value to be established on the basis of cellular details and of expression of immunohistochemical marker

*e) Bioptic sampling*

Samples will be collected, from surgical specimens of tumors arriving fresh (unfixed) from the Surgical Theatre to the Pathology labs. Samples will be obtained from pathological areas of the following organs: Breast, Colon, Uterus, Prostate and Lung. No specific risks for specimens' collection are foreseen.

*f) Possible interactions with concomitant medical treatments*

Not applicable.

*g) Steps that will be taken to control or mitigate the risks, based on the risk-analysis assessment*

Not applicable.

*h) Risk-to-benefit rationale.*

The performance of Glyoxal Acid-Free (GAF) is given by the fact that represents a novel tissue fixative by investigating morphological preservation and diagnostic value to be established on the basis of cellular details and of expression of immunohistochemical marker.

The use of Glyoxal Acid-Free (GAF) should not represent any particular risk. However,

*This document is confidential and is to be distributed for review only to investigators, consultants, study staff, and applicable Independent Ethics Committees National Competent Authorities or Institutional Review Boards. The contents of this document shall not be disclosed to others without written authorization from Sponsor.*

possible side effects shall be considered, like the above mentioned Anticipated adverse device effects to any of the device's ingredient, misuse of the device that might compromise the positive effect of the device when correctly used.

#### 4.6 MONITORING PLAN

The data was recorded in an eCRF. The Investigator entered data and performed corrections as per GCP requirements.
The study monitor (CRA) contacted and visited the investigational site at study initiation, throughout the study and after the study completion to perform the site closure visit. CRA verified the various study records: eCRF, ISF and source data (source data is any information in original records and certified copies of original records on clinical findings, observations or other activities in a study necessary for the reconstruction and evaluation of the study). Source data are contained in source documents – in fully respect of subject's confidentiality - in order to fulfill both the sponsor's responsibility in assuring the proper conduct of the study regarding protocol and GCP adherence and the completeness and accuracy of the data recorded on the eCRF.

The Investigator and/or study team members were expected to be available during the monitoring visits, to answer questions and to provide any missing information. Onsite Monitoring visits were not always possible, due to COVID-19 restrictions. The monitoring visits scheduling was performed as reported below:

| | **Site 1** | **Site 2** | **Site 3** |
|---|---|---|---|
| **MONITORING VISITS** | **Date** | **Date** | **Date** |
| Site Initiation Visit (SIV) | 30 JAN 2020 | 25 JUN 2020 | 19-20 FEB 2020 |
| Monitoring Visit (MV) 1 | 02 JUL 2021 | Not performed | 19 OCT 2021 |

*This document is confidential and is to be distributed for review only to investigators, consultants, study staff, and applicable Independent Ethics Committees National Competent Authorities or Institutional Review Boards. The contents of this document shall not be disclosed to others without written authorization from Sponsor.*

| | | | |
|---|---|---|---|
| | | due to COVID-19 pandemic | |
| Close-Out Visit (COV) | Planned in Q1 2022 | Planned in Q1 2022 | Planned in Q1 2022 |

*This document is confidential and is to be distributed for review only to investigators, consultants, study staff, and applicable Independent Ethics Committees National Competent Authorities or Institutional Review Boards. The contents of this document shall not be disclosed to others without written authorization from Sponsor.*

### 5 RESULTS

#### 5.1 DISPOSITION OF SLIDES

During the study 200 slides were collected and consequently included in the analysis*.

All the slides were included in accordance with SAP in each analysis as "all analyzed slides".

There are exactly 100 slides collected for each used fixative (GAF and PBF). In particular, 42 (21.0%) slides were collected in Manchester and 68 (34.0%) in Barcelona; the majority of slides were collected in Candiolo (45.0%). (Table 1)

**Table 1**. Analyzed slides by center and by used fixative

| Center | | Slides (N=200) |
|---|---|---|
| Manchester | N (%) | 42 (21.0) |
| Barcelona | N (%) | 68 (34.0) |
| Candiolo | N (%) | 90 (45.0) |
| Fixative | | |
| GAF | N (%) | 100 (50.0) |
| PBF | N (%) | 100 (50.0) |

*__Note__: From each case (100 histological samples), 2 paraffin-embedded tissue blocks were obtained. From the tissue blocks, 5 Slides (1 H&E + 4 IHC) were produced, which were scanned and presented (a total of 1000 slides) for reading by Reviewers.*

#### 5.2 PROTOCOL VIOLATIONS

No Protocol violations were recorded during the entire study.

*This document is confidential and is to be distributed for review only to investigators, consultants, study staff, and applicable Independent Ethics Committees National Competent Authorities or Institutional Review Boards. The contents of this document shall not be disclosed to others without written authorization from Sponsor.*

#### 5.3 EFFICACY ANALYSIS

##### *5.3.1 Primary efficacy analysis*

The primary endpoint of the study was the evaluation of morphological preservation and diagnostic value questions answered by Central Pathology Reviewer in terms of the total score obtained in GAF group compared with total score calculated in PBF group.

The assumption of normality distribution of total score data is violated (Shapiro-Wilk Test, $P < 0.0001$), so in this analysis, the difference between the two fixative groups was tested using a non-parametric approach.

The mean of total score in GAF fixative group is 3.7 ± 0.5 while in PBF fixative group is 3.9 ± 0.3.

However, in terms of median value we observe similar results between fixative groups, a median value of 4.0 (IQR: 3.5-4.0) with a minimum value of 2.0 and a maximum of 4.0 and a median value of 4.0 (IQR: 4.0-4.0) with a minimum value of 3.0 and a maximum of 4.0 were observed in GAF fixative group and in PBF fixative group, respectively. (Table 2)

Applying Wilcoxon Signed-Rank test to test the non-inferiority (-2.0 of non-inferiority margin), P-value is less than 0.001, therefore the null hypothesis of inferiority can be rejected and the non-inferiority of Glyoxal Acid-Free Fixative to the reference Phosphate buffered Formalin could be considered achieved.

Moreover, the estimate of the mean difference between fixative groups (GAF and PBF) was -0.20 with a 95% CI from -0.30 to -0.09 and the non-inferiority margin is not included.

The primary efficacy analysis was performed also by organs.

Out of 90 slides collected from breast, the total score mean value for GAF fixative group is 3.7 ± 0.6 and it is 3.9 ± 0.3 for PBF fixative group. As for the overall primary efficacy analysis, the median is 4.0 (IQR: 4.0-4.0) in both fixative groups. In this case the mean difference between fixative groups is -0.2 ± 0.6. (Table 3)

Applying Wilcoxon Signed-Rank test again to test the non-inferiority exclusively in breast slides (-2.0 of non-inferiority margin), P-value is less than 0.001, therefore the null hypothesis

*This document is confidential and is to be distributed for review only to investigators, consultants, study staff, and applicable Independent Ethics Committees National Competent Authorities or Institutional Review Boards. The contents of this document shall not be disclosed to others without written authorization from Sponsor.*

of inferiority can be rejected. Also, the non-inferiority of Glyoxal Acid-Free Fixative to the reference Phosphate buffered Formalin could be considered achieved in the breast subgroup.

Moreover, the estimate of the mean difference between fixative groups in breast slide (GAF and PBF) was -0.24 with a 95% CI from -0.42 to -0.07 and the non-inferiority margin is not included.

Regarding the 52 slides collected from colon, the total score mean value for GAF fixative group is 3.7 ± 0.5 and 3.9 ± 0.3 for PBF fixative group, similar to the results obtained from breast slides. The median value is 4.0 (IQR: 3.0-4.0) in GAF fixative group and it is 4.0 (IQR: 4.0-4.0) in PBF fixative group. (Table 3)

Applying Wilcoxon Signed-Rank test again to test the non-inferiority, P-value is less than 0.001, therefore the null hypothesis of inferiority can be rejected and also the non-inferiority of Glyoxal Acid-Free Fixative to the reference Phosphate buffered Formalin could be considered achieved in the subgroup of colon slides.

Moreover, the estimate of the mean difference between fixative groups in colon slide (GAF and PBF) was -0.19 with a 95% CI from -0.39 to 0.01 and the non-inferiority margin is not included.

All 16 slides collected from lung received high score from Central Pathology Reviewer. The total score mean value for GAF fixative group is 3.9 ± 0.4 and it is 4.0 ± 0.0 for PBF fixative group. As for the overall primary efficacy analysis, the median is 4.0 (IQR: 4.0-4.0) in both fixative groups. In this case the mean difference between fixative groups is -0.1 ± 0.4. (Table 3)

Applying Wilcoxon Signed-Rank test again to test the non-inferiority exclusively in lung slides (-2.0 of non-inferiority margin), P-value is less than 0.001, therefore the null hypothesis of inferiority can be rejected. Also, the non-inferiority of Glyoxal Acid-Free Fixative to the reference Phosphate buffered Formalin could be considered achieved in the lung subgroup.

*This document is confidential and is to be distributed for review only to investigators, consultants, study staff, and applicable Independent Ethics Committees National Competent Authorities or Institutional Review Boards. The contents of this document shall not be disclosed to others without written authorization from Sponsor.*

Moreover, the estimate of the mean difference between fixative groups in lung slide (GAF and PBF) was -0.12 with a 95% CI from -0.42 to -0.17 and the non-inferiority margin is not included.

Regarding the 22 slides collected from endometrium, the total score mean value for GAF fixative group is 3.8 ± 0.4 and 3.9 ± 0.3 for PBF fixative group, similar to the results obtained from breast slides. The median value is 4.0 (IQR: 4.0-4.0) in both fixative groups. (Table 3)

Applying Wilcoxon Signed-Rank test again to test the non-inferiority, P-value is less than 0.001, therefore the null hypothesis of inferiority can be rejected and also the non-inferiority of Glyoxal Acid-Free Fixative to the reference Phosphate buffered Formalin could be considered achieved in the subgroup of endometrium slides.

Moreover, the estimate of the mean difference between fixative groups in colon slide (GAF and PBF) was -0.09 with a 95% CI from -0.45 to 0.27 and the non-inferiority margin is not included.

For the prostate 20 slides were collected, 10 per fixative group; both received similar score from Central Pathology Reviewer. In particular, total score mean value for GAF fixative group is 3.7 ± 0.5 and 3.9 ± 0.3 for PBF fixative group, similar to the results obtained from breast slides. The median value is 4.0 (IQR: 4.0-4.0) in both fixative groups. (Table 3)

Applying Wilcoxon Signed-Rank test again to test the non-inferiority, P-value is less than 0.001, therefore the null hypothesis of inferiority can be rejected and also the non-inferiority of Glyoxal Acid-Free Fixative to the reference Phosphate buffered Formalin could be considered achieved in the subgroup of prostate slides.

Moreover, the estimate of the mean difference between fixative groups in colon slide (GAF and PBF) was -0.20 with a 95% CI from -0.65 to 0.25 and the non-inferiority margin is not included.

*This document is confidential and is to be distributed for review only to investigators, consultants, study staff, and applicable Independent Ethics Committees National Competent Authorities or Institutional Review Boards. The contents of this document shall not be disclosed to others without written authorization from Sponsor.*

**Table 2**. Primary Efficacy Endpoint

| Central reviewer Total Score | Fixative | | | |
|---|---|---|---|---|
| **Overall (N=200)** | **GAF** | **PBF** | **Difference between fixative** | **P [1]** |
| **N** | 100 | 100 | 100 | <0.001 |
| **Mean ± SD** | 3.7 ± 0.5 | 3.9 ± 0.3 | -0.2 ± 0.5 | |
| **Median (IQR)** | 4.0 (3.5-4.0) | 4.0 (4.0-4.0) | 0.0 (0.0-0.0) | |
| **(Min-Max)** | 2.0-4.0 | 3.0-4.0 | -2.0-1.0 | |

SD: Standard Deviation

IQR: Interquartile Range

P[1]: P-value from Wilcoxon Signed-Rank to test non-inferiority of GAF

Notes:

a) Difference between fixative is defined as GAF total score – PBF total score

**Table 3**. Primary Efficacy Endpoint by organs

| Central reviewer Total Score by organ | Fixative | | | |
|---|---|---|---|---|
| **Organ** | **GAF** | **PBF** | **Difference between fixative** | **P [1]** |
| **Breast (N=90)** | | | | |
| **N** | 45 | 45 | 45 | <0.001 |
| **Mean ± SD** | 3.7 ± 0.6 | 3.9 ± 0.3 | -0.2 ± 0.6 | |
| **Median (IQR)** | 4.0 (4.0-4.0) | 4.0 (4.0-4.0) | 0.0 (0.0-0.0) | |

*This document is confidential and is to be distributed for review only to investigators, consultants, study staff, and applicable Independent Ethics Committees National Competent Authorities or Institutional Review Boards. The contents of this document shall not be disclosed to others without written authorization from Sponsor.*

| | | | | |
|---|---|---|---|---|
| **(Min-Max)** | 2.0-4.0 | 2.0-4.0 | -2.0-1.0 | |
| **Colon (N=52)** | | | | <0.001 |
| **N** | 26 | 26 | 26 | |
| **Mean ± SD** | 3.7 ± 0.5 | 3.9 ± 0.3 | -0.2 ± 0.5 | |
| **Median (IQR)** | 4.0 (3.0-4.0) | 4.0 (4.0-4.0) | 0.0 (-1.0-0.0) | |
| **(Min-Max)** | 3.0-4.0 | 3.0-4.0 | -1.0-1.0 | |
| **Lung (N=16)** | | | | <0.001 |
| **N** | 8 | 8 | 8 | |
| **Mean ± SD** | 3.9 ± 0.4 | 4.0 ± 0.0 | -0.1 ± 0.4 | |
| **Median (IQR)** | 4.0 (4.0-4.0) | 4.0 (4.0-4.0) | 0.0 (0.0-0.0) | |
| **(Min-Max)** | 3.0-4.0 | 4.0-4.0 | -1.0-0.0 | |
| **Endometrium (N=22)** | | | | <0.001 |
| **N** | 11 | 11 | 11 | |
| **Mean ± SD** | 3.8 ± 0.4 | 3.9 ± 0.3 | -0.1 ± 0.5 | |
| **Median (IQR)** | 4.0 (4.0-4.0) | 4.0 (4.0-4.0) | 0.0 (0.0-0.0) | |
| **(Min-Max)** | 3.0-4.0 | 3.0-4.0 | -1.0-1.0 | |
| **Prostate (N=20)** | | | | <0.001 |
| **N** | 10 | 10 | 10 | |
| **Mean ± SD** | 3.7 ± 0.5 | 3.9 ± 0.3 | -0.2 ± 0.6 | |
| **Median (IQR)** | 4.0 (3.0-4.0) | 4.0 (4.0-4.0) | 0.0 (0.0-0.0) | |
| **(Min-Max)** | 3.0-4.0 | 4.0-4.0 | -1.0-1.0 | |

SD: Standard Deviation

IQR: Interquartile Range

P[1]: P-value from Wilcoxon Signed-Rank to test non-inferiority of GAF

Notes:

a) Difference between fixative is defined as GAF total score – PBF total score

*This document is confidential and is to be distributed for review only to investigators, consultants, study staff, and applicable Independent Ethics Committees National Competent Authorities or Institutional Review Boards. The contents of this document shall not be disclosed to others without written authorization from Sponsor.*

##### 5.3.2 *Analysis of secondary analysis*

The first secondary endpoint of the study was the evaluation of morphological preservation and diagnostic value questions answered by local center pathologists in terms of the total score obtained in GAF group compared with total score calculated in PBF group.

The assumption of normality distribution of total score data is violated (Shapiro-Wilk Test, $P < 0.0001$), so in this analysis, the difference between the two fixative groups was tested using a non-parametric approach.

The mean of total score in GAF fixative group is 3.8 ± 0.5 while in PBF fixative group is 4.0 ± 0.1.

However, in terms of median value we observe similar results between fixative groups, a median value of 4.0 (IQR: 4.0-4.0) with a minimum value of 1.0 and a maximum of 4.0 and a median value of 4.0 (IQR: 4.0-4.0) with a minimum value of 3.0 and a maximum of 4.0 were observed in GAF fixative group and in PBF fixative group, respectively. (Table 2)

The only case that scored 1 point was a GAF prostate case analyzed in Manchester (slide "06" regarding prostate). The same case received a score of 3 points from the central reviewer. (Listing 3b, Listing 4)

In general, the scores obtained by the local pathologist are very similar to those received by the central reviewer. Applying Wilcoxon Signed-Rank test to test the non-inferiority (-2.0 of non-inferiority margin), P-value is less than 0.001, therefore the null hypothesis of inferiority can be rejected and the non-inferiority of Glyoxal Acid-Free Fixative to the reference Phosphate buffered Formalin could be considered achieved.

Moreover, the estimate of the mean difference between fixative groups (GAF and PBF) was -0.21 with a 95% CI from -0.31 to -0.11 and the non-inferiority margin is not included.

The second secondary endpoint of the study was the evaluation of Pathologist's satisfaction on local centers. The question concerning satisfaction was only asked for GAF cases and the score went from 1 (not satisfied) to 10 (totally satisfied). The higher mean satisfaction was

*This document is confidential and is to be distributed for review only to investigators, consultants, study staff, and applicable Independent Ethics Committees National Competent Authorities or Institutional Review Boards. The contents of this document shall not be disclosed to others without written authorization from Sponsor.*

recorded in Barcelona (9.8 ± 0.7), with a median of 10 and a minimum of 7 points. (Table 5) The overall mean satisfaction can be considered positive. In particular, overall local center pathologist mean was 9.2 ± 1.1, with a median value of 9.6 points (IQR: 8.7-10.0), a minimum value of 5.4 and a maximum of 10.0. (Table 5)

Regarding the question asked to local centers "Do you consider that the preparations obtained on the same case with the two fixatives have the same performance?", 18 (18.0%) cases were answered negatively, while all the remaining 82 (82.0%) cases were answered "Yes". (Table 5)

The fifth question asked at the centers therefore cannot provide the same detail and accuracy as the score composed of the four questions concerning structural preservation of the tissue, preservation of the nuclei, preservation of the cytoplasm and diagnostic value of these preparations.

**Table 4**. Total score from four questions evaluated by local reviewers

| Local center pathologist Total Score | Fixative | | | |
|---|---|---|---|---|
| **Overall (N=200)** | **GAF** | **PBF** | **Difference between fixative** | **P [1]** |
| **N** | 100 | 100 | 100 | <0.001 |
| **Mean ± SD** | 3.8 ± 0.5 | 4.0 ± 0.1 | -0.2 ± 0.5 | |
| **Median (IQR)** | 4.0 (3.0-4.0) | 4.0 (4.0-4.0) | 0.0 (0.0-0.0) | |
| **(Min-Max)** | 1.0-4.0 | 3.0-4.0 | -3.0-1.0 | |

SD: Standard Deviation
IQR: Interquartile Range
P[1]: P-value from Wilcoxon Signed-Rank to test non-inferiority of GAF
Notes:
a) Difference between fixative is defined as GAF total score – PBF total score

*This document is confidential and is to be distributed for review only to investigators, consultants, study staff, and applicable Independent Ethics Committees National Competent Authorities or Institutional Review Boards. The contents of this document shall not be disclosed to others without written authorization from Sponsor.*

**Table 5**. Pathologist satisfaction

| Local center pathologist satisfaction | Study center | | | | |
|---|---|---|---|---|---|
| | Manchester | Barcelona | Candiolo | Overall | P [1] |
| N | 21 | 34 | 45 | 100 | <0.001 |
| Mean ± SD | 8.8 ± 1.6 | 9.8 ± 0.7 | 9.0 ± 0.7 | 9.2 ± 1.1 | |
| Median (IQR) | 9.6 (8.3-10.0) | 10.0 (10.0-10.0) | 9.0 (8.5-9.5) | 9.6 (8.7-10.0) | |
| (Min-Max) | 5.4-10.0 | 7.0-10.0 | 7.0-10.0 | 5.4-10.0 | |
| Fifth question | | | | | |
| Yes - N (%) | 11 (52.4) | 28 (82.4) | 43 (95.5) | 82 (82.0) | |
| No - N (%) | 10 (47.6) | 6 (17.7) | 2 (4.5) | 18 (18.0) | |

SD: Standard Deviation
IQR: Interquartile Range
P[1]: P-value from Wilcoxon Signed-Rank to test non-inferiority of GAF
Notes:

a) Difference between fixative is defined as GAF total score – PBF total score

##### 5.3.3 *Analysis of other variables*

No analysis of other variables has been planned for this study.

*This document is confidential and is to be distributed for review only to investigators, consultants, study staff, and applicable Independent Ethics Committees National Competent Authorities or Institutional Review Boards. The contents of this document shall not be disclosed to others without written authorization from Sponsor.*

### 6 DISCUSSION AND OVERALL CONCLUSIONS

#### 6.1 CLINICAL SAFETY AND PERFORMANCE RESULTS

At the end of the study, all primary and secondary endpoints were performed.

In total, 200 slides were considered in the analysis, in particular 100 slides (50%) were performed on GAF and 100 slides (50%) were performed on PBF.

The performance results confirmed the non-inferiority of GAF respect to PBF as highlighted by the previous interim analysis. Therefore, the GAF has the same ability of tissue fixation without toxicity and carcinogenic activity.

In this study, a classic safety evaluation (adverse events, vital signs and physical examination) was not planned, because there was not a direct involvement of subject or patient. However, as mentioned above, Glyoxal was proposed in 1943 (10) as a fixative alternative to formalin since it is a simple di-aldehyde. As reported by Harke & Hoeffler (11) glyoxal does not appear to evaporate from solution. Indeed, the reported Henry's law constant of ≤3.38 × 10– 4Pa m3/mol (12) indicates that glyoxal is essentially non-volatile with regard to the aqueous phase. Glyoxal is not classifiable as a human carcinogen (13), nevertheless it use may cause some adverse reactions such as irritation of skin and eyes (13).

For the secondary analysis, the performance evaluations for the local laboratories was considered. Also, in this case the results confirmed the non-inferiority of GAF respect to PBF and in terms of median value similar results between fixative groups were observed.

The overall mean satisfaction of the local laboratories can be considered positive. The mean value, considering a score scale from 1 (not satisfied) to 10 (totally satisfied), was 9.2 ± 1.1. The only two insufficient scores were recorded by the Manchester laboratory (5.4 and 5.5 for a prostate and a endometrium slide respectively) even if all the parameter of performance evaluated (structural preservation of the tissue, preservation of the nuclei, preservation of the cytoplasm and diagnostic value) were considered valid.

*This document is confidential and is to be distributed for review only to investigators, consultants, study staff, and applicable Independent Ethics Committees National Competent Authorities or Institutional Review Boards. The contents of this document shall not be disclosed to others without written authorization from Sponsor.*

#### 6.2 ASSESSMENT OF RISKS AND BENEFITS

No clinical benefits are expected as the aim of the currently proposed study is to confirm in a large sample of histological specimens, obtained from different tissues, that an acid-free form of glyoxal (GAF) represents a novel tissue fixative by investigating morphological preservation and diagnostic value to be established on the basis of cellular details and of expression of immunohistochemical marker

As expected, the use of Glyoxal Acid-Free (GAF) seems not represent any particular risk. However, possible side effects were considered, like the above mentioned Anticipated adverse device effects to any of the device's ingredient, misuse of the device that might compromise the positive effect of the device when correctly used.

#### 6.3 CLINICAL RELEVANCE AND IMPORTANCE OF THE RESULTS

Confirming the non-inferiority of GAF respect to PBF, the data of the trial highlight the capability of the investigational device to ensure the structural preservation of the: tissue, nuclei, cytoplasm and diagnostic value of the preparations (for the sections of all organs tested). These results are consistent with the rational/justification of the study and confirmed the satisfaction of local laboratories.

#### 6.4 SPECIFIC BENEFITS OR SPECIAL PRECAUTIONS

As described above, considering the toxicity of formalin, environmental authorities are increasingly concerned for the objective toxicity of this volatile reagent, so that a banning of formalin from 2016 has been proposed in the European union. This has been stated by the ec regulation n.605/2014 (4) that modifies the ec regulation n.1272/2008 (5) defining formalin as a carcinogen (category 1b/2) and mutagen. This regulation should heavily impact on diagnostic pathology methods and procedures, even if so far the main reaction to this significant issue is limited to adoption of protective procedures, designed to prevent excessive exposure of pathology workers to formaldehyde vapors.

*This document is confidential and is to be distributed for review only to investigators, consultants, study staff, and applicable Independent Ethics Committees National Competent Authorities or Institutional Review Boards. The contents of this document shall not be disclosed to others without written authorization from Sponsor.*

At the end of this Clinical Performance Study, the data of this report support GAF could be considered an effective tool as well PBF for the diagnostic procedures for the tumor specimens considered (breast, prostate, colon, endometrium, and lung), but with lower safety concerns than PBF itself.

On the other hand, considering the potential advantages related to the use of GAF, since the fixation time for the tissue specimen was set as 3 h. for GAF, 6 h. for PBF, the results imply that GAF is not only safer, but also faster than PBF, parity of results being considered, potentially speeding up the histological staining procedures.

#### 6.5 IMPLICATIONS FOR FUTURE STUDIES

Even if from literature it is recognized the safety of GAF, on the other hand no data on the safety were collected during this Clinical Performance Study. In future studies, it could be useful to also consider some safety items (e.g., regarding the potential misuse of the IVD) to maintain updated the product vigilance. On the other hand, it could be interesting to expand the plethora of other tumor specimens to be analyzed. In this way, the promising result of this Clinical Performance Study could be verified also in the diagnostic process of other tumors kinds.

#### 6.6 LIMITATIONS OF THE STUDY

A limitation of the study is potentially due to the size of the tissue specimens here investigated, since only small biopsies (and not large specimens were considered. An additional consideration is concerning the questionnaire on the satisfaction of local laboratory, that only allowed binary answers. It would have been useful to know the justification of the negative scores on the performance parameters (structural preservation of the tissue, preservation of the nuclei, preservation of the cytoplasm and diagnostic value).

*This document is confidential and is to be distributed for review only to investigators, consultants, study staff, and applicable Independent Ethics Committees National Competent Authorities or Institutional Review Boards. The contents of this document shall not be disclosed to others without written authorization from Sponsor.*

Also the results obtained with the question: "Do you consider that the preparations obtained on the same case with the two fixatives have the same performance?" can bring some consideration. Eighteen (18.0%) cases were answered negatively. This evaluation was observed both with high and low satisfaction scores. In this case could have been useful to clarify when this discrepancy was due to a considered superiority of the investigational product or not.

The above remarks offer prospects for future focused investigations.

*This document is confidential and is to be distributed for review only to investigators, consultants, study staff, and applicable Independent Ethics Committees National Competent Authorities or Institutional Review Boards. The contents of this document shall not be disclosed to others without written authorization from Sponsor.*

### 7 LISTING

**Listing 1**. All analyzed and fixed slides

| Slides | Center | Evaluated organ | Fixative |
|---|---|---|---|
| 01 | Barcelona | Colon | GAF |
| 01 | Barcelona | Colon | PBF |
| 01 | Barcelona | Lung | GAF |
| 01 | Barcelona | Lung | PBF |
| 01 | Manchester | Endometrium | GAF |
| 01 | Manchester | Endometrium | PBF |
| 02 | Candiolo | Breast | GAF |
| 02 | Candiolo | Breast | PBF |
| 02 | Barcelona | Colon | GAF |
| 02 | Barcelona | Colon | PBF |
| 02 | Barcelona | Lung | GAF |
| 02 | Barcelona | Lung | PBF |
| 02 | Manchester | Endometrium | GAF |
| 02 | Manchester | Endometrium | PBF |
| 03 | Candiolo | Breast | GAF |
| 03 | Candiolo | Breast | PBF |
| 03 | Barcelona | Colon | GAF |
| 03 | Barcelona | Colon | PBF |
| 03 | Barcelona | Lung | GAF |
| 03 | Barcelona | Lung | PBF |
| 04 | Candiolo | Breast | GAF |
| 04 | Candiolo | Breast | PBF |
| 04 | Barcelona | Colon | GAF |
| 04 | Barcelona | Colon | PBF |
| 04 | Barcelona | Lung | GAF |

*This document is confidential and is to be distributed for review only to investigators, consultants, study staff, and applicable Independent Ethics Committees National Competent Authorities or Institutional Review Boards. The contents of this document shall not be disclosed to others without written authorization from Sponsor.*

| Slides | Center | Evaluated organ | Fixative |
|---|---|---|---|
| 04 | Barcelona | Lung | PBF |
| 04 | Manchester | Endometrium | GAF |
| 04 | Manchester | Endometrium | PBF |
| 04 | Manchester | Prostate | GAF |
| 04 | Manchester | Prostate | PBF |
| 05 | Barcelona | Colon | GAF |
| 05 | Barcelona | Colon | PBF |
| 05 | Barcelona | Lung | GAF |
| 05 | Barcelona | Lung | PBF |
| 05 | Manchester | Endometrium | GAF |
| 05 | Manchester | Endometrium | PBF |
| 06 | Candiolo | Breast | GAF |
| 06 | Candiolo | Breast | PBF |
| 06 | Barcelona | Colon | GAF |
| 06 | Barcelona | Colon | PBF |
| 06 | Barcelona | Lung | GAF |
| 06 | Barcelona | Lung | PBF |
| 06 | Manchester | Prostate | GAF |
| 06 | Manchester | Prostate | PBF |
| 07 | Candiolo | Breast | GAF |
| 07 | Candiolo | Breast | PBF |
| 07 | Barcelona | Colon | GAF |
| 07 | Barcelona | Colon | PBF |
| 07 | Barcelona | Lung | GAF |
| 07 | Barcelona | Lung | PBF |
| 07 | Manchester | Endometrium | GAF |
| 07 | Manchester | Endometrium | PBF |

*This document is confidential and is to be distributed for review only to investigators, consultants, study staff, and applicable Independent Ethics Committees National Competent Authorities or Institutional Review Boards. The contents of this document shall not be disclosed to others without written authorization from Sponsor.*

| Slides | Center | Evaluated organ | Fixative |
|---|---|---|---|
| 07 | Manchester | Prostate | GAF |
| 07 | Manchester | Prostate | PBF |
| 08 | Candiolo | Breast | GAF |
| 08 | Candiolo | Breast | PBF |
| 08 | Barcelona | Colon | GAF |
| 08 | Barcelona | Colon | PBF |
| 08 | Barcelona | Lung | GAF |
| 08 | Barcelona | Lung | PBF |
| 08 | Manchester | Endometrium | GAF |
| 08 | Manchester | Endometrium | PBF |
| 08 | Manchester | Prostate | GAF |
| 08 | Manchester | Prostate | PBF |
| 09 | Candiolo | Breast | GAF |
| 09 | Candiolo | Breast | PBF |
| 09 | Barcelona | Colon | GAF |
| 09 | Barcelona | Colon | PBF |
| 09 | Manchester | Endometrium | GAF |
| 09 | Manchester | Endometrium | PBF |
| 09 | Manchester | Prostate | GAF |
| 09 | Manchester | Prostate | PBF |
| 10 | Barcelona | Colon | GAF |
| 10 | Barcelona | Colon | PBF |
| 10 | Manchester | Endometrium | GAF |
| 10 | Manchester | Endometrium | PBF |
| 10 | Manchester | Prostate | GAF |
| 10 | Manchester | Prostate | PBF |
| 11 | Candiolo | Breast | GAF |

*This document is confidential and is to be distributed for review only to investigators, consultants, study staff, and applicable Independent Ethics Committees National Competent Authorities or Institutional Review Boards. The contents of this document shall not be disclosed to others without written authorization from Sponsor.*

| Slides | Center | Evaluated organ | Fixative |
|---|---|---|---|
| 11 | Candiolo | Breast | PBF |
| 11 | Manchester | Endometrium | GAF |
| 11 | Manchester | Endometrium | PBF |
| 11 | Manchester | Prostate | GAF |
| 11 | Manchester | Prostate | PBF |
| 12 | Candiolo | Breast | GAF |
| 12 | Candiolo | Breast | PBF |
| 12 | Barcelona | Colon | GAF |
| 12 | Barcelona | Colon | PBF |
| 12 | Manchester | Endometrium | GAF |
| 12 | Manchester | Endometrium | PBF |
| 12 | Manchester | Prostate | GAF |
| 12 | Manchester | Prostate | PBF |
| 13 | Candiolo | Breast | GAF |
| 13 | Candiolo | Breast | PBF |
| 13 | Barcelona | Colon | GAF |
| 13 | Barcelona | Colon | PBF |
| 13 | Manchester | Endometrium | GAF |
| 13 | Manchester | Endometrium | PBF |
| 13 | Manchester | Prostate | GAF |
| 13 | Manchester | Prostate | PBF |
| 14 | Candiolo | Breast | GAF |
| 14 | Candiolo | Breast | PBF |
| 14 | Barcelona | Colon | GAF |
| 14 | Barcelona | Colon | PBF |
| 14 | Manchester | Prostate | GAF |
| 14 | Manchester | Prostate | PBF |

*This document is confidential and is to be distributed for review only to investigators, consultants, study staff, and applicable Independent Ethics Committees National Competent Authorities or Institutional Review Boards. The contents of this document shall not be disclosed to others without written authorization from Sponsor.*

| Slides | Center | Evaluated organ | Fixative |
|---|---|---|---|
| 15 | Candiolo | Breast | GAF |
| 15 | Candiolo | Breast | PBF |
| 15 | Barcelona | Colon | GAF |
| 15 | Barcelona | Colon | PBF |
| 16 | Candiolo | Breast | GAF |
| 16 | Candiolo | Breast | PBF |
| 16 | Barcelona | Colon | GAF |
| 16 | Barcelona | Colon | PBF |
| 17 | Candiolo | Breast | GAF |
| 17 | Candiolo | Breast | PBF |
| 17 | Barcelona | Colon | GAF |
| 17 | Barcelona | Colon | PBF |
| 18 | Barcelona | Colon | GAF |
| 18 | Barcelona | Colon | PBF |
| 19 | Candiolo | Breast | GAF |
| 19 | Candiolo | Breast | PBF |
| 19 | Barcelona | Colon | GAF |
| 19 | Barcelona | Colon | PBF |
| 20 | Candiolo | Breast | GAF |
| 20 | Candiolo | Breast | PBF |
| 20 | Barcelona | Colon | GAF |
| 20 | Barcelona | Colon | PBF |
| 21 | Candiolo | Breast | GAF |
| 21 | Candiolo | Breast | PBF |
| 21 | Barcelona | Colon | GAF |
| 21 | Barcelona | Colon | PBF |
| 22 | Candiolo | Breast | GAF |

*This document is confidential and is to be distributed for review only to investigators, consultants, study staff, and applicable Independent Ethics Committees National Competent Authorities or Institutional Review Boards. The contents of this document shall not be disclosed to others without written authorization from Sponsor.*

| Slides | Center | Evaluated organ | Fixative |
|---|---|---|---|
| 22 | Candiolo | Breast | PBF |
| 22 | Barcelona | Colon | GAF |
| 22 | Barcelona | Colon | PBF |
| 23 | Candiolo | Breast | GAF |
| 23 | Candiolo | Breast | PBF |
| 23 | Barcelona | Colon | GAF |
| 23 | Barcelona | Colon | PBF |
| 24 | Candiolo | Breast | GAF |
| 24 | Candiolo | Breast | PBF |
| 24 | Barcelona | Colon | GAF |
| 24 | Barcelona | Colon | PBF |
| 25 | Candiolo | Breast | GAF |
| 25 | Candiolo | Breast | PBF |
| 25 | Barcelona | Colon | GAF |
| 25 | Barcelona | Colon | PBF |
| 26 | Candiolo | Breast | GAF |
| 26 | Candiolo | Breast | PBF |
| 26 | Barcelona | Colon | GAF |
| 26 | Barcelona | Colon | PBF |
| 27 | Candiolo | Breast | GAF |
| 27 | Candiolo | Breast | PBF |
| 27 | Barcelona | Colon | GAF |
| 27 | Barcelona | Colon | PBF |
| 28 | Candiolo | Breast | GAF |
| 28 | Candiolo | Breast | PBF |
| 29 | Candiolo | Breast | GAF |
| 29 | Candiolo | Breast | PBF |

*This document is confidential and is to be distributed for review only to investigators, consultants, study staff, and applicable Independent Ethics Committees National Competent Authorities or Institutional Review Boards. The contents of this document shall not be disclosed to others without written authorization from Sponsor.*

| Slides | Center | Evaluated organ | Fixative |
|---|---|---|---|
| 30 | Candiolo | Breast | GAF |
| 30 | Candiolo | Breast | PBF |
| 31 | Candiolo | Breast | GAF |
| 31 | Candiolo | Breast | PBF |
| 33 | Candiolo | Breast | GAF |
| 33 | Candiolo | Breast | PBF |
| 34 | Candiolo | Breast | GAF |
| 34 | Candiolo | Breast | PBF |
| 35 | Candiolo | Breast | GAF |
| 35 | Candiolo | Breast | PBF |
| 36 | Candiolo | Breast | GAF |
| 36 | Candiolo | Breast | PBF |
| 37 | Candiolo | Breast | GAF |
| 37 | Candiolo | Breast | PBF |
| 38 | Candiolo | Breast | GAF |
| 38 | Candiolo | Breast | PBF |
| 40 | Candiolo | Breast | GAF |
| 40 | Candiolo | Breast | PBF |
| 41 | Candiolo | Breast | GAF |
| 41 | Candiolo | Breast | PBF |
| 42 | Candiolo | Breast | GAF |
| 42 | Candiolo | Breast | PBF |
| 43 | Candiolo | Breast | GAF |
| 43 | Candiolo | Breast | PBF |
| 44 | Candiolo | Breast | GAF |
| 44 | Candiolo | Breast | PBF |
| 45 | Candiolo | Breast | GAF |

*This document is confidential and is to be distributed for review only to investigators, consultants, study staff, and applicable Independent Ethics Committees National Competent Authorities or Institutional Review Boards. The contents of this document shall not be disclosed to others without written authorization from Sponsor.*

| Slides | Center | Evaluated organ | Fixative |
|---|---|---|---|
| 45 | Candiolo | Breast | PBF |
| 46 | Candiolo | Breast | GAF |
| 46 | Candiolo | Breast | PBF |
| 47 | Candiolo | Breast | GAF |
| 47 | Candiolo | Breast | PBF |
| 48 | Candiolo | Breast | GAF |
| 48 | Candiolo | Breast | PBF |
| 50 | Candiolo | Breast | GAF |
| 50 | Candiolo | Breast | PBF |
| 51 | Candiolo | Breast | GAF |
| 51 | Candiolo | Breast | PBF |
| 52 | Candiolo | Breast | GAF |
| 52 | Candiolo | Breast | PBF |

**Listing 2a**. Primary Efficacy Endpoint

| Slides | Fixative | Evaluated organ | How do you regard the structural preservation of the tissue | How do you regard the preservation of the nuclei | How do you regard the preservation of the cytoplasm | How do you regard the diagnostic value of these preparations |
|---|---|---|---|---|---|---|
| 01 | GAF | Colon | Valid | Valid | Valid | Valid |
| 01 | PBF | Colon | Valid | Valid | Valid | Valid |
| 01 | GAF | Lung | Valid | Valid | Valid | Valid |
| 01 | PBF | Lung | Valid | Valid | Valid | Valid |
| 01 | GAF | Endometrium | Valid | Valid | Valid | Valid |
| 01 | PBF | Endometrium | Valid | Valid | Valid | Valid |
| 02 | GAF | Breast | Valid | Valid | Valid | Valid |
| 02 | PBF | Breast | Valid | Valid | Valid | Valid |

*This document is confidential and is to be distributed for review only to investigators, consultants, study staff, and applicable Independent Ethics Committees National Competent Authorities or Institutional Review Boards. The contents of this document shall not be disclosed to others without written authorization from Sponsor.*

| Slides | Fixative | Evaluated organ | How do you regard the structural preservation of the tissue | How do you regard the preservation of the nuclei | How do you regard the preservation of the cytoplasm | How do you regard the diagnostic value of these preparations |
|---|---|---|---|---|---|---|
| 02 | GAF | Colon | Valid | Valid | Valid | Valid |
| 02 | PBF | Colon | Valid | Valid | Valid | Valid |
| 02 | GAF | Lung | Valid | Valid | Valid | Valid |
| 02 | PBF | Lung | Valid | Valid | Valid | Valid |
| 02 | GAF | Endometrium | Valid | Valid | Valid | Valid |
| 02 | PBF | Endometrium | Valid | Valid | Valid | Valid |
| 03 | GAF | Breast | Valid | Valid | Valid | Valid |
| 03 | PBF | Breast | Valid | Valid | Valid | Valid |
| 03 | GAF | Colon | Valid | Valid | Valid | Valid |
| 03 | PBF | Colon | Valid | Valid | Valid | Valid |
| 03 | GAF | Lung | Valid | Valid | Valid | Valid |
| 03 | PBF | Lung | Valid | Valid | Valid | Valid |
| 04 | GAF | Breast | Valid | Valid | Valid | Valid |
| 04 | PBF | Breast | Valid | Valid | Valid | Valid |
| 04 | GAF | Colon | Valid | Valid | Valid | Valid |
| 04 | PBF | Colon | Valid | Valid | Valid | Valid |
| 04 | GAF | Lung | Valid | Valid | Valid | Valid |
| 04 | PBF | Lung | Valid | Valid | Valid | Valid |
| 04 | GAF | Endometrium | Valid | Valid | Valid | Valid |
| 04 | PBF | Endometrium | Valid | Valid | Valid | Valid |
| 04 | GAF | Prostate | Valid | Valid | Valid | Valid |
| 04 | PBF | Prostate | Valid | Valid | Valid | Valid |
| 05 | GAF | Colon | Valid | Valid | Valid | Valid |
| 05 | PBF | Colon | Valid | Valid | Valid | Valid |
| 05 | GAF | Lung | Valid | Valid | Valid | Valid |

*This document is confidential and is to be distributed for review only to investigators, consultants, study staff, and applicable Independent Ethics Committees National Competent Authorities or Institutional Review Boards. The contents of this document shall not be disclosed to others without written authorization from Sponsor.*

| Slides | Fixative | Evaluated organ | How do you regard the structural preservation of the tissue | How do you regard the preservation of the nuclei | How do you regard the preservation of the cytoplasm | How do you regard the diagnostic value of these preparations |
|---|---|---|---|---|---|---|
| 05 | PBF | Lung | Valid | Valid | Valid | Valid |
| 05 | GAF | Endometrium | Valid | Valid | Valid | Valid |
| 05 | PBF | Endometrium | Valid | Valid | Valid | Valid |
| 06 | GAF | Breast | Valid | Valid | Valid | Valid |
| 06 | PBF | Breast | Valid | Valid | Valid | Valid |
| 06 | GAF | Colon | Valid | Valid | Valid | Valid |
| 06 | PBF | Colon | Valid | Valid | Valid | Valid |
| 06 | GAF | Lung | Valid | Valid | Valid | Valid |
| 06 | PBF | Lung | Valid | Valid | Valid | Valid |
| 06 | GAF | Prostate | Invalid | Valid | Valid | Valid |
| 06 | PBF | Prostate | Valid | Valid | Valid | Valid |
| 07 | GAF | Breast | Invalid | Valid | Valid | Valid |
| 07 | PBF | Breast | Valid | Valid | Valid | Valid |
| 07 | GAF | Colon | Valid | Valid | Valid | Valid |
| 07 | PBF | Colon | Valid | Valid | Valid | Valid |
| 07 | GAF | Lung | Valid | Valid | Valid | Valid |
| 07 | PBF | Lung | Valid | Valid | Valid | Valid |
| 07 | GAF | Endometrium | Valid | Valid | Valid | Invalid |
| 07 | PBF | Endometrium | Valid | Valid | Valid | Valid |
| 07 | GAF | Prostate | Valid | Valid | Valid | Valid |
| 07 | PBF | Prostate | Valid | Valid | Valid | Valid |
| 08 | GAF | Breast | Invalid | Valid | Valid | Valid |
| 08 | PBF | Breast | Valid | Valid | Valid | Valid |
| 08 | GAF | Colon | Valid | Valid | Valid | Valid |
| 08 | PBF | Colon | Valid | Valid | Valid | Invalid |

*This document is confidential and is to be distributed for review only to investigators, consultants, study staff, and applicable Independent Ethics Committees National Competent Authorities or Institutional Review Boards. The contents of this document shall not be disclosed to others without written authorization from Sponsor.*

| Slides | Fixative | Evaluated organ | How do you regard the structural preservation of the tissue | How do you regard the preservation of the nuclei | How do you regard the preservation of the cytoplasm | How do you regard the diagnostic value of these preparations |
|---|---|---|---|---|---|---|
| 08 | GAF | Lung | Invalid | Valid | Valid | Valid |
| 08 | PBF | Lung | Valid | Valid | Valid | Valid |
| 08 | GAF | Endometrium | Valid | Valid | Valid | Valid |
| 08 | PBF | Endometrium | Valid | Valid | Valid | Valid |
| 08 | GAF | Prostate | Valid | Valid | Valid | Valid |
| 08 | PBF | Prostate | Valid | Valid | Valid | Valid |
| 09 | GAF | Breast | Invalid | Valid | Valid | Valid |
| 09 | PBF | Breast | Valid | Valid | Valid | Valid |
| 09 | GAF | Colon | Valid | Valid | Valid | Invalid |
| 09 | PBF | Colon | Valid | Valid | Valid | Invalid |
| 09 | GAF | Endometrium | Valid | Valid | Valid | Valid |
| 09 | PBF | Endometrium | Valid | Valid | Valid | Valid |
| 09 | GAF | Prostate | Valid | Valid | Valid | Valid |
| 09 | PBF | Prostate | Valid | Valid | Valid | Valid |
| 10 | GAF | Colon | Valid | Valid | Valid | Invalid |
| 10 | PBF | Colon | Valid | Valid | Valid | Valid |
| 10 | GAF | Endometrium | Valid | Valid | Valid | Valid |
| 10 | PBF | Endometrium | Valid | Valid | Valid | Valid |
| 10 | GAF | Prostate | Valid | Valid | Valid | Valid |
| 10 | PBF | Prostate | Valid | Valid | Valid | Valid |
| 11 | GAF | Breast | Valid | Valid | Valid | Valid |
| 11 | PBF | Breast | Valid | Valid | Valid | Valid |
| 11 | GAF | Endometrium | Invalid | Valid | Valid | Valid |
| 11 | PBF | Endometrium | Valid | Valid | Valid | Valid |
| 11 | GAF | Prostate | Valid | Valid | Valid | Valid |

*This document is confidential and is to be distributed for review only to investigators, consultants, study staff, and applicable Independent Ethics Committees National Competent Authorities or Institutional Review Boards. The contents of this document shall not be disclosed to others without written authorization from Sponsor.*

| Slides | Fixative | Evaluated organ | How do you regard the structural preservation of the tissue | How do you regard the preservation of the nuclei | How do you regard the preservation of the cytoplasm | How do you regard the diagnostic value of these preparations |
|---|---|---|---|---|---|---|
| 11 | PBF | Prostate | Valid | Valid | Valid | Invalid |
| 12 | GAF | Breast | Valid | Valid | Valid | Valid |
| 12 | PBF | Breast | Valid | Valid | Valid | Invalid |
| 12 | GAF | Colon | Valid | Valid | Valid | Valid |
| 12 | PBF | Colon | Valid | Valid | Valid | Valid |
| 12 | GAF | Endometrium | Valid | Valid | Valid | Valid |
| 12 | PBF | Endometrium | Valid | Valid | Valid | Valid |
| 12 | GAF | Prostate | Invalid | Valid | Valid | Valid |
| 12 | PBF | Prostate | Valid | Valid | Valid | Valid |
| 13 | GAF | Breast | Invalid | Invalid | Valid | Valid |
| 13 | PBF | Breast | Valid | Valid | Valid | Valid |
| 13 | GAF | Colon | Valid | Valid | Valid | Invalid |
| 13 | PBF | Colon | Valid | Valid | Valid | Valid |
| 13 | GAF | Endometrium | Valid | Valid | Valid | Valid |
| 13 | PBF | Endometrium | Valid | Valid | Valid | Invalid |
| 13 | GAF | Prostate | Valid | Valid | Valid | Valid |
| 13 | PBF | Prostate | Valid | Valid | Valid | Valid |
| 14 | GAF | Breast | Valid | Valid | Valid | Valid |
| 14 | PBF | Breast | Valid | Valid | Valid | Valid |
| 14 | GAF | Colon | Valid | Valid | Valid | Valid |
| 14 | PBF | Colon | Valid | Valid | Valid | Valid |
| 14 | GAF | Prostate | Invalid | Valid | Valid | Valid |
| 14 | PBF | Prostate | Valid | Valid | Valid | Valid |
| 15 | GAF | Breast | Valid | Valid | Valid | Valid |
| 15 | PBF | Breast | Valid | Valid | Valid | Valid |

*This document is confidential and is to be distributed for review only to investigators, consultants, study staff, and applicable Independent Ethics Committees National Competent Authorities or Institutional Review Boards. The contents of this document shall not be disclosed to others without written authorization from Sponsor.*

| Slides | Fixative | Evaluated organ | How do you regard the structural preservation of the tissue | How do you regard the preservation of the nuclei | How do you regard the preservation of the cytoplasm | How do you regard the diagnostic value of these preparations |
|---|---|---|---|---|---|---|
| 15 | GAF | Colon | Valid | Valid | Valid | Valid |
| 15 | PBF | Colon | Valid | Valid | Valid | Valid |
| 16 | GAF | Breast | Valid | Valid | Valid | Valid |
| 16 | PBF | Breast | Valid | Valid | Valid | Valid |
| 16 | GAF | Colon | Valid | Valid | Valid | Valid |
| 16 | PBF | Colon | Valid | Valid | Valid | Valid |
| 17 | GAF | Breast | Valid | Valid | Valid | Valid |
| 17 | PBF | Breast | Valid | Valid | Valid | Valid |
| 17 | GAF | Colon | Valid | Valid | Valid | Valid |
| 17 | PBF | Colon | Valid | Valid | Valid | Valid |
| 18 | GAF | Colon | Valid | Valid | Valid | Valid |
| 18 | PBF | Colon | Valid | Valid | Valid | Valid |
| 19 | GAF | Breast | Valid | Valid | Valid | Valid |
| 19 | PBF | Breast | Valid | Valid | Valid | Valid |
| 19 | GAF | Colon | Valid | Valid | Valid | Valid |
| 19 | PBF | Colon | Valid | Valid | Valid | Valid |
| 20 | GAF | Breast | Valid | Valid | Valid | Valid |
| 20 | PBF | Breast | Valid | Valid | Valid | Valid |
| 20 | GAF | Colon | Invalid | Valid | Valid | Valid |
| 20 | PBF | Colon | Valid | Valid | Valid | Valid |
| 21 | GAF | Breast | Valid | Valid | Valid | Invalid |
| 21 | PBF | Breast | Valid | Valid | Valid | Valid |
| 21 | GAF | Colon | Invalid | Valid | Valid | Valid |
| 21 | PBF | Colon | Valid | Valid | Valid | Valid |
| 22 | GAF | Breast | Valid | Valid | Valid | Invalid |

*This document is confidential and is to be distributed for review only to investigators, consultants, study staff, and applicable Independent Ethics Committees National Competent Authorities or Institutional Review Boards. The contents of this document shall not be disclosed to others without written authorization from Sponsor.*

| Slides | Fixative | Evaluated organ | How do you regard the structural preservation of the tissue | How do you regard the preservation of the nuclei | How do you regard the preservation of the cytoplasm | How do you regard the diagnostic value of these preparations |
|---|---|---|---|---|---|---|
| 22 | PBF | Breast | Valid | Valid | Valid | Valid |
| 22 | GAF | Colon | Invalid | Valid | Valid | Valid |
| 22 | PBF | Colon | Valid | Valid | Valid | Valid |
| 23 | GAF | Breast | Valid | Valid | Valid | Valid |
| 23 | PBF | Breast | Valid | Valid | Valid | Valid |
| 23 | GAF | Colon | Invalid | Valid | Valid | Valid |
| 23 | PBF | Colon | Valid | Valid | Valid | Valid |
| 24 | GAF | Breast | Invalid | Valid | Valid | Valid |
| 24 | PBF | Breast | Valid | Valid | Valid | Valid |
| 24 | GAF | Colon | Valid | Valid | Valid | Invalid |
| 24 | PBF | Colon | Valid | Valid | Valid | Invalid |
| 25 | GAF | Breast | Valid | Valid | Valid | Valid |
| 25 | PBF | Breast | Valid | Valid | Valid | Valid |
| 25 | GAF | Colon | Valid | Valid | Valid | Valid |
| 25 | PBF | Colon | Valid | Valid | Valid | Valid |
| 26 | GAF | Breast | Valid | Valid | Valid | Valid |
| 26 | PBF | Breast | Valid | Valid | Valid | Valid |
| 26 | GAF | Colon | Valid | Valid | Valid | Valid |
| 26 | PBF | Colon | Valid | Valid | Valid | Valid |
| 27 | GAF | Breast | Valid | Valid | Valid | Valid |
| 27 | PBF | Breast | Valid | Valid | Valid | Valid |
| 27 | GAF | Colon | Valid | Valid | Valid | Valid |
| 27 | PBF | Colon | Valid | Valid | Valid | Valid |
| 28 | GAF | Breast | Valid | Valid | Valid | Valid |

*This document is confidential and is to be distributed for review only to investigators, consultants, study staff, and applicable Independent Ethics Committees National Competent Authorities or Institutional Review Boards. The contents of this document shall not be disclosed to others without written authorization from Sponsor.*

| Slides | Fixative | Evaluated organ | How do you regard the structural preservation of the tissue | How do you regard the preservation of the nuclei | How do you regard the preservation of the cytoplasm | How do you regard the diagnostic value of these preparations |
|---|---|---|---|---|---|---|
| 28 | PBF | Breast | Valid | Valid | Valid | Valid |
| 29 | GAF | Breast | Valid | Valid | Valid | Valid |
| 29 | PBF | Breast | Valid | Valid | Valid | Valid |
| 30 | GAF | Breast | Valid | Valid | Valid | Valid |
| 30 | PBF | Breast | Valid | Valid | Valid | Valid |
| 31 | GAF | Breast | Valid | Valid | Valid | Valid |
| 31 | PBF | Breast | Valid | Valid | Valid | Valid |
| 33 | GAF | Breast | Valid | Valid | Valid | Valid |
| 33 | PBF | Breast | Valid | Valid | Valid | Valid |
| 34 | GAF | Breast | Invalid | Valid | Valid | Valid |
| 34 | PBF | Breast | Invalid | Valid | Valid | Valid |
| 35 | GAF | Breast | Valid | Valid | Valid | Valid |
| 35 | PBF | Breast | Valid | Valid | Valid | Valid |
| 36 | GAF | Breast | Valid | Valid | Valid | Valid |
| 36 | PBF | Breast | Valid | Valid | Valid | Valid |
| 37 | GAF | Breast | Valid | Valid | Valid | Valid |
| 37 | PBF | Breast | Valid | Valid | Valid | Valid |
| 38 | GAF | Breast | Valid | Valid | Valid | Valid |
| 38 | PBF | Breast | Valid | Valid | Valid | Valid |
| 40 | GAF | Breast | Valid | Valid | Valid | Valid |
| 40 | PBF | Breast | Valid | Valid | Valid | Valid |
| 41 | GAF | Breast | Invalid | Valid | Valid | Invalid |
| 41 | PBF | Breast | Valid | Valid | Valid | Valid |
| 42 | GAF | Breast | Valid | Valid | Valid | Valid |

*This document is confidential and is to be distributed for review only to investigators, consultants, study staff, and applicable Independent Ethics Committees National Competent Authorities or Institutional Review Boards. The contents of this document shall not be disclosed to others without written authorization from Sponsor.*

| Slides | Fixative | Evaluated organ | How do you regard the structural preservation of the tissue | How do you regard the preservation of the nuclei | How do you regard the preservation of the cytoplasm | How do you regard the diagnostic value of these preparations |
|---|---|---|---|---|---|---|
| 42 | PBF | Breast | Valid | Valid | Valid | Valid |
| 43 | GAF | Breast | Valid | Valid | Valid | Valid |
| 43 | PBF | Breast | Valid | Valid | Valid | Valid |
| 44 | GAF | Breast | Valid | Valid | Valid | Valid |
| 44 | PBF | Breast | Valid | Valid | Valid | Valid |
| 45 | GAF | Breast | Valid | Valid | Valid | Valid |
| 45 | PBF | Breast | Valid | Valid | Valid | Valid |
| 46 | GAF | Breast | Valid | Valid | Valid | Valid |
| 46 | PBF | Breast | Valid | Valid | Valid | Valid |
| 47 | GAF | Breast | Invalid | Valid | Valid | Invalid |
| 47 | PBF | Breast | Valid | Valid | Valid | Invalid |
| 48 | GAF | Breast | Valid | Valid | Valid | Valid |
| 48 | PBF | Breast | Valid | Valid | Valid | Valid |
| 50 | GAF | Breast | Valid | Valid | Valid | Valid |
| 50 | PBF | Breast | Valid | Valid | Valid | Valid |
| 51 | GAF | Breast | Valid | Valid | Valid | Invalid |
| 51 | PBF | Breast | Valid | Valid | Valid | Valid |
| 52 | GAF | Breast | Valid | Valid | Valid | Valid |
| 52 | PBF | Breast | Valid | Valid | Valid | Valid |

**Listing 2b**. Primary Efficacy Endpoint

| Slides | Fixative | Evaluated organ | Central reviewer Total Score |
|---|---|---|---|
| 01 | GAF | Colon | 4 |

| Slides | Fixative | Evaluated organ | Central reviewer Total Score |
|---|---|---|---|
| 01 | PBF | Colon | 4 |
| 01 | GAF | Lung | 4 |
| 01 | PBF | Lung | 4 |
| 01 | GAF | Endometrium | 4 |
| 01 | PBF | Endometrium | 4 |
| 02 | GAF | Breast | 4 |
| 02 | PBF | Breast | 4 |
| 02 | GAF | Colon | 4 |
| 02 | PBF | Colon | 4 |
| 02 | GAF | Lung | 4 |
| 02 | PBF | Lung | 4 |
| 02 | GAF | Endometrium | 4 |
| 02 | PBF | Endometrium | 4 |
| 03 | GAF | Breast | 4 |
| 03 | PBF | Breast | 4 |
| 03 | GAF | Colon | 4 |
| 03 | PBF | Colon | 4 |
| 03 | GAF | Lung | 4 |
| 03 | PBF | Lung | 4 |
| 04 | GAF | Breast | 4 |
| 04 | PBF | Breast | 4 |
| 04 | GAF | Colon | 4 |
| 04 | PBF | Colon | 4 |
| 04 | GAF | Lung | 4 |
| 04 | PBF | Lung | 4 |
| 04 | GAF | Endometrium | 4 |
| 04 | PBF | Endometrium | 4 |

*This document is confidential and is to be distributed for review only to investigators, consultants, study staff, and applicable Independent Ethics Committees National Competent Authorities or Institutional Review Boards. The contents of this document shall not be disclosed to others without written authorization from Sponsor.*

| Slides | Fixative | Evaluated organ | Central reviewer Total Score |
|---|---|---|---|
| 04 | GAF | Prostate | 4 |
| 04 | PBF | Prostate | 4 |
| 05 | GAF | Colon | 4 |
| 05 | PBF | Colon | 4 |
| 05 | GAF | Lung | 4 |
| 05 | PBF | Lung | 4 |
| 05 | GAF | Endometrium | 4 |
| 05 | PBF | Endometrium | 4 |
| 06 | GAF | Breast | 4 |
| 06 | PBF | Breast | 4 |
| 06 | GAF | Colon | 4 |
| 06 | PBF | Colon | 4 |
| 06 | GAF | Lung | 4 |
| 06 | PBF | Lung | 4 |
| 06 | GAF | Prostate | 3 |
| 06 | PBF | Prostate | 4 |
| 07 | GAF | Breast | 3 |
| 07 | PBF | Breast | 4 |
| 07 | GAF | Colon | 4 |
| 07 | PBF | Colon | 4 |
| 07 | GAF | Lung | 4 |
| 07 | PBF | Lung | 4 |
| 07 | GAF | Endometrium | 3 |
| 07 | PBF | Endometrium | 4 |
| 07 | GAF | Prostate | 4 |
| 07 | PBF | Prostate | 4 |
| 08 | GAF | Breast | 3 |

*This document is confidential and is to be distributed for review only to investigators, consultants, study staff, and applicable Independent Ethics Committees National Competent Authorities or Institutional Review Boards. The contents of this document shall not be disclosed to others without written authorization from Sponsor.*

| Slides | Fixative | Evaluated organ | Central reviewer Total Score |
|---|---|---|---|
| 08 | PBF | Breast | 4 |
| 08 | GAF | Colon | 4 |
| 08 | PBF | Colon | 3 |
| 08 | GAF | Lung | 3 |
| 08 | PBF | Lung | 4 |
| 08 | GAF | Endometrium | 4 |
| 08 | PBF | Endometrium | 4 |
| 08 | GAF | Prostate | 4 |
| 08 | PBF | Prostate | 4 |
| 09 | GAF | Breast | 3 |
| 09 | PBF | Breast | 4 |
| 09 | GAF | Colon | 3 |
| 09 | PBF | Colon | 3 |
| 09 | GAF | Endometrium | 4 |
| 09 | PBF | Endometrium | 4 |
| 09 | GAF | Prostate | 4 |
| 09 | PBF | Prostate | 4 |
| 10 | GAF | Colon | 3 |
| 10 | PBF | Colon | 4 |
| 10 | GAF | Endometrium | 4 |
| 10 | PBF | Endometrium | 4 |
| 10 | GAF | Prostate | 4 |
| 10 | PBF | Prostate | 4 |
| 11 | GAF | Breast | 4 |
| 11 | PBF | Breast | 4 |
| 11 | GAF | Endometrium | 3 |
| 11 | PBF | Endometrium | 4 |

*This document is confidential and is to be distributed for review only to investigators, consultants, study staff, and applicable Independent Ethics Committees National Competent Authorities or Institutional Review Boards. The contents of this document shall not be disclosed to others without written authorization from Sponsor.*

| Slides | Fixative | Evaluated organ | Central reviewer Total Score |
|---|---|---|---|
| 11 | GAF | Prostate | 4 |
| 11 | PBF | Prostate | 3 |
| 12 | GAF | Breast | 4 |
| 12 | PBF | Breast | 3 |
| 12 | GAF | Colon | 4 |
| 12 | PBF | Colon | 4 |
| 12 | GAF | Endometrium | 4 |
| 12 | PBF | Endometrium | 4 |
| 12 | GAF | Prostate | 3 |
| 12 | PBF | Prostate | 4 |
| 13 | GAF | Breast | 2 |
| 13 | PBF | Breast | 4 |
| 13 | GAF | Colon | 3 |
| 13 | PBF | Colon | 4 |
| 13 | GAF | Endometrium | 4 |
| 13 | PBF | Endometrium | 3 |
| 13 | GAF | Prostate | 4 |
| 13 | PBF | Prostate | 4 |
| 14 | GAF | Breast | 4 |
| 14 | PBF | Breast | 4 |
| 14 | GAF | Colon | 4 |
| 14 | PBF | Colon | 4 |
| 14 | GAF | Prostate | 3 |
| 14 | PBF | Prostate | 4 |
| 15 | GAF | Breast | 4 |
| 15 | PBF | Breast | 4 |
| 15 | GAF | Colon | 4 |

*This document is confidential and is to be distributed for review only to investigators, consultants, study staff, and applicable Independent Ethics Committees National Competent Authorities or Institutional Review Boards. The contents of this document shall not be disclosed to others without written authorization from Sponsor.*

| Slides | Fixative | Evaluated organ | Central reviewer Total Score |
|---|---|---|---|
| 15 | PBF | Colon | 4 |
| 16 | GAF | Breast | 4 |
| 16 | PBF | Breast | 4 |
| 16 | GAF | Colon | 4 |
| 16 | PBF | Colon | 4 |
| 17 | GAF | Breast | 4 |
| 17 | PBF | Breast | 4 |
| 17 | GAF | Colon | 4 |
| 17 | PBF | Colon | 4 |
| 18 | GAF | Colon | 4 |
| 18 | PBF | Colon | 4 |
| 19 | GAF | Breast | 4 |
| 19 | PBF | Breast | 4 |
| 19 | GAF | Colon | 4 |
| 19 | PBF | Colon | 4 |
| 20 | GAF | Breast | 4 |
| 20 | PBF | Breast | 4 |
| 20 | GAF | Colon | 3 |
| 20 | PBF | Colon | 4 |
| 21 | GAF | Breast | 3 |
| 21 | PBF | Breast | 4 |
| 21 | GAF | Colon | 3 |
| 21 | PBF | Colon | 4 |
| 22 | GAF | Breast | 3 |
| 22 | PBF | Breast | 4 |
| 22 | GAF | Colon | 3 |
| 22 | PBF | Colon | 4 |

*This document is confidential and is to be distributed for review only to investigators, consultants, study staff, and applicable Independent Ethics Committees National Competent Authorities or Institutional Review Boards. The contents of this document shall not be disclosed to others without written authorization from Sponsor.*

| Slides | Fixative | Evaluated organ | Central reviewer Total Score |
|---|---|---|---|
| 23 | GAF | Breast | 4 |
| 23 | PBF | Breast | 4 |
| 23 | GAF | Colon | 3 |
| 23 | PBF | Colon | 4 |
| 24 | GAF | Breast | 3 |
| 24 | PBF | Breast | 4 |
| 24 | GAF | Colon | 3 |
| 24 | PBF | Colon | 3 |
| 25 | GAF | Breast | 4 |
| 25 | PBF | Breast | 4 |
| 25 | GAF | Colon | 4 |
| 25 | PBF | Colon | 4 |
| 26 | GAF | Breast | 4 |
| 26 | PBF | Breast | 4 |
| 26 | GAF | Colon | 4 |
| 26 | PBF | Colon | 4 |
| 27 | GAF | Breast | 4 |
| 27 | PBF | Breast | 4 |
| 27 | GAF | Colon | 4 |
| 27 | PBF | Colon | 4 |
| 28 | GAF | Breast | 4 |
| 28 | PBF | Breast | 4 |
| 29 | GAF | Breast | 4 |
| 29 | PBF | Breast | 4 |
| 30 | GAF | Breast | 4 |
| 30 | PBF | Breast | 4 |

*This document is confidential and is to be distributed for review only to investigators, consultants, study staff, and applicable Independent Ethics Committees National Competent Authorities or Institutional Review Boards. The contents of this document shall not be disclosed to others without written authorization from Sponsor.*

| Slides | Fixative | Evaluated organ | Central reviewer Total Score |
|---|---|---|---|
| 31 | GAF | Breast | 4 |
| 31 | PBF | Breast | 4 |
| 33 | GAF | Breast | 4 |
| 33 | PBF | Breast | 4 |
| 34 | GAF | Breast | 3 |
| 34 | PBF | Breast | 3 |
| 35 | GAF | Breast | 4 |
| 35 | PBF | Breast | 4 |
| 36 | GAF | Breast | 4 |
| 36 | PBF | Breast | 4 |
| 37 | GAF | Breast | 4 |
| 37 | PBF | Breast | 4 |
| 38 | GAF | Breast | 4 |
| 38 | PBF | Breast | 4 |
| 40 | GAF | Breast | 4 |
| 40 | PBF | Breast | 4 |
| 41 | GAF | Breast | 2 |
| 41 | PBF | Breast | 4 |
| 42 | GAF | Breast | 4 |
| 42 | PBF | Breast | 4 |
| 43 | GAF | Breast | 4 |
| 43 | PBF | Breast | 4 |
| 44 | GAF | Breast | 4 |
| 44 | PBF | Breast | 4 |
| 45 | GAF | Breast | 4 |
| 45 | PBF | Breast | 4 |

*This document is confidential and is to be distributed for review only to investigators, consultants, study staff, and applicable Independent Ethics Committees National Competent Authorities or Institutional Review Boards. The contents of this document shall not be disclosed to others without written authorization from Sponsor.*

| Slides | Fixative | Evaluated organ | Central reviewer Total Score |
|---|---|---|---|
| 46 | GAF | Breast | 4 |
| 46 | PBF | Breast | 4 |
| 47 | GAF | Breast | 2 |
| 47 | PBF | Breast | 3 |
| 48 | GAF | Breast | 4 |
| 48 | PBF | Breast | 4 |
| 50 | GAF | Breast | 4 |
| 50 | PBF | Breast | 4 |
| 51 | GAF | Breast | 3 |
| 51 | PBF | Breast | 4 |
| 52 | GAF | Breast | 4 |
| 52 | PBF | Breast | 4 |

**Listing 3a**. Secondary Efficacy Endpoints

| Slides | Fixative | Evaluated organ | How do you regard the structural preservation of the tissue | How do you regard the preservation of the nuclei | How do you regard the preservation of the cytoplasm | How do you regard the diagnostic value of these preparations |
|---|---|---|---|---|---|---|
| 01 | GAF | Colon | Valid | Valid | Valid | Valid |
| 01 | PBF | Colon | Valid | Valid | Valid | Valid |
| 01 | GAF | Lung | Valid | Valid | Invalid | Valid |
| 01 | PBF | Lung | Valid | Valid | Valid | Valid |
| 01 | GAF | Endometrium | Valid | Valid | Valid | Valid |
| 01 | PBF | Endometrium | Valid | Valid | Valid | Valid |
| 02 | GAF | Breast | Valid | Valid | Valid | Valid |
| 02 | PBF | Breast | Valid | Valid | Valid | Valid |
| 02 | GAF | Colon | Valid | Valid | Valid | Valid |

*This document is confidential and is to be distributed for review only to investigators, consultants, study staff, and applicable Independent Ethics Committees National Competent Authorities or Institutional Review Boards. The contents of this document shall not be disclosed to others without written authorization from Sponsor.*

| Slides | Fixative | Evaluated organ | How do you regard the structural preservation of the tissue | How do you regard the preservation of the nuclei | How do you regard the preservation of the cytoplasm | How do you regard the diagnostic value of these preparations |
|---|---|---|---|---|---|---|
| 02 | PBF | Colon | Valid | Valid | Valid | Valid |
| 02 | GAF | Lung | Valid | Valid | Valid | Valid |
| 02 | PBF | Lung | Valid | Valid | Valid | Valid |
| 02 | GAF | Endometrium | Valid | Valid | Valid | Valid |
| 02 | PBF | Endometrium | Valid | Valid | Valid | Valid |
| 03 | GAF | Breast | Valid | Valid | Valid | Valid |
| 03 | PBF | Breast | Valid | Valid | Valid | Valid |
| 03 | GAF | Colon | Valid | Valid | Valid | Valid |
| 03 | PBF | Colon | Valid | Valid | Valid | Valid |
| 03 | GAF | Lung | Valid | Valid | Valid | Valid |
| 03 | PBF | Lung | Valid | Valid | Valid | Valid |
| 04 | GAF | Breast | Valid | Valid | Valid | Valid |
| 04 | PBF | Breast | Valid | Valid | Valid | Valid |
| 04 | GAF | Colon | Valid | Valid | Valid | Valid |
| 04 | PBF | Colon | Valid | Valid | Valid | Valid |
| 04 | GAF | Lung | Valid | Valid | Valid | Valid |
| 04 | PBF | Lung | Valid | Valid | Valid | Valid |
| 04 | GAF | Endometrium | Valid | Valid | Valid | Valid |
| 04 | PBF | Endometrium | Valid | Valid | Valid | Valid |
| 04 | GAF | Prostate | Valid | Valid | Valid | Valid |
| 04 | PBF | Prostate | Valid | Valid | Valid | Valid |
| 05 | GAF | Colon | Valid | Valid | Valid | Valid |
| 05 | PBF | Colon | Valid | Valid | Valid | Valid |
| 05 | GAF | Lung | Valid | Valid | Valid | Valid |
| 05 | PBF | Lung | Valid | Valid | Valid | Valid |

*This document is confidential and is to be distributed for review only to investigators, consultants, study staff, and applicable Independent Ethics Committees National Competent Authorities or Institutional Review Boards. The contents of this document shall not be disclosed to others without written authorization from Sponsor.*

| Slides | Fixative | Evaluated organ | How do you regard the structural preservation of the tissue | How do you regard the preservation of the nuclei | How do you regard the preservation of the cytoplasm | How do you regard the diagnostic value of these preparations |
|---|---|---|---|---|---|---|
| 05 | GAF | Endometrium | Valid | Valid | Valid | Valid |
| 05 | PBF | Endometrium | Valid | Valid | Valid | Valid |
| 06 | GAF | Breast | Valid | Valid | Valid | Valid |
| 06 | PBF | Breast | Valid | Valid | Valid | Valid |
| 06 | GAF | Colon | Valid | Valid | Valid | Valid |
| 06 | PBF | Colon | Valid | Valid | Valid | Valid |
| 06 | GAF | Lung | Valid | Valid | Valid | Valid |
| 06 | PBF | Lung | Valid | Valid | Valid | Valid |
| 06 | GAF | Prostate | Invalid | Invalid | Invalid | Valid |
| 06 | PBF | Prostate | Valid | Valid | Valid | Valid |
| 07 | GAF | Breast | Valid | Valid | Valid | Valid |
| 07 | PBF | Breast | Valid | Valid | Valid | Valid |
| 07 | GAF | Colon | Valid | Valid | Valid | Valid |
| 07 | PBF | Colon | Valid | Valid | Valid | Valid |
| 07 | GAF | Lung | Valid | Valid | Valid | Valid |
| 07 | PBF | Lung | Valid | Valid | Valid | Valid |
| 07 | GAF | Endometrium | Valid | Valid | Valid | Invalid |
| 07 | PBF | Endometrium | Valid | Valid | Valid | Valid |
| 07 | GAF | Prostate | Valid | Invalid | Valid | Valid |
| 07 | PBF | Prostate | Valid | Valid | Valid | Valid |
| 08 | GAF | Breast | Valid | Valid | Valid | Valid |
| 08 | PBF | Breast | Valid | Valid | Valid | Valid |
| 08 | GAF | Colon | Valid | Valid | Valid | Valid |
| 08 | PBF | Colon | Valid | Valid | Valid | Valid |
| 08 | GAF | Lung | Valid | Valid | Valid | Valid |

*This document is confidential and is to be distributed for review only to investigators, consultants, study staff, and applicable Independent Ethics Committees National Competent Authorities or Institutional Review Boards. The contents of this document shall not be disclosed to others without written authorization from Sponsor.*

| Slides | Fixative | Evaluated organ | How do you regard the structural preservation of the tissue | How do you regard the preservation of the nuclei | How do you regard the preservation of the cytoplasm | How do you regard the diagnostic value of these preparations |
|---|---|---|---|---|---|---|
| 08 | PBF | Lung | Valid | Valid | Valid | Valid |
| 08 | GAF | Endometrium | Valid | Valid | Valid | Valid |
| 08 | PBF | Endometrium | Valid | Valid | Valid | Valid |
| 08 | GAF | Prostate | Valid | Invalid | Valid | Valid |
| 08 | PBF | Prostate | Valid | Valid | Valid | Valid |
| 09 | GAF | Breast | Valid | Valid | Valid | Valid |
| 09 | PBF | Breast | Valid | Valid | Valid | Valid |
| 09 | GAF | Colon | Valid | Valid | Valid | Invalid |
| 09 | PBF | Colon | Valid | Valid | Valid | Valid |
| 09 | GAF | Endometrium | Valid | Valid | Valid | Valid |
| 09 | PBF | Endometrium | Valid | Valid | Valid | Valid |
| 09 | GAF | Prostate | Valid | Invalid | Valid | Valid |
| 09 | PBF | Prostate | Valid | Valid | Valid | Valid |
| 10 | GAF | Colon | Valid | Valid | Valid | Valid |
| 10 | PBF | Colon | Valid | Valid | Valid | Valid |
| 10 | GAF | Endometrium | Valid | Valid | Valid | Valid |
| 10 | PBF | Endometrium | Valid | Valid | Valid | Valid |
| 10 | GAF | Prostate | Valid | Invalid | Valid | Valid |
| 10 | PBF | Prostate | Valid | Valid | Valid | Valid |
| 11 | GAF | Breast | Valid | Valid | Valid | Valid |
| 11 | PBF | Breast | Valid | Valid | Valid | Valid |
| 11 | GAF | Endometrium | Valid | Valid | Valid | Invalid |
| 11 | PBF | Endometrium | Valid | Valid | Valid | Valid |
| 11 | GAF | Prostate | Valid | Invalid | Valid | Valid |
| 11 | PBF | Prostate | Valid | Valid | Valid | Valid |

*This document is confidential and is to be distributed for review only to investigators, consultants, study staff, and applicable Independent Ethics Committees National Competent Authorities or Institutional Review Boards. The contents of this document shall not be disclosed to others without written authorization from Sponsor.*

| Slides | Fixative | Evaluated organ | How do you regard the structural preservation of the tissue | How do you regard the preservation of the nuclei | How do you regard the preservation of the cytoplasm | How do you regard the diagnostic value of these preparations |
|---|---|---|---|---|---|---|
| 12 | GAF | Breast | Valid | Valid | Valid | Valid |
| 12 | PBF | Breast | Valid | Valid | Valid | Valid |
| 12 | GAF | Colon | Valid | Valid | Valid | Valid |
| 12 | PBF | Colon | Valid | Valid | Valid | Valid |
| 12 | GAF | Endometrium | Valid | Valid | Valid | Invalid |
| 12 | PBF | Endometrium | Valid | Valid | Valid | Valid |
| 12 | GAF | Prostate | Valid | Invalid | Valid | Valid |
| 12 | PBF | Prostate | Valid | Valid | Valid | Valid |
| 13 | GAF | Breast | Valid | Valid | Valid | Valid |
| 13 | PBF | Breast | Valid | Valid | Valid | Valid |
| 13 | GAF | Colon | Valid | Valid | Valid | Valid |
| 13 | PBF | Colon | Valid | Valid | Valid | Valid |
| 13 | GAF | Endometrium | Valid | Valid | Valid | Valid |
| 13 | PBF | Endometrium | Valid | Valid | Valid | Valid |
| 13 | GAF | Prostate | Valid | Invalid | Valid | Valid |
| 13 | PBF | Prostate | Valid | Valid | Valid | Valid |
| 14 | GAF | Breast | Valid | Valid | Valid | Valid |
| 14 | PBF | Breast | Valid | Valid | Valid | Valid |
| 14 | GAF | Colon | Valid | Valid | Valid | Valid |
| 14 | PBF | Colon | Valid | Valid | Valid | Valid |
| 14 | GAF | Prostate | Valid | Invalid | Valid | Valid |
| 14 | PBF | Prostate | Valid | Valid | Valid | Valid |
| 15 | GAF | Breast | Valid | Valid | Valid | Valid |
| 15 | PBF | Breast | Valid | Valid | Valid | Valid |
| 15 | GAF | Colon | Valid | Valid | Valid | Valid |

*This document is confidential and is to be distributed for review only to investigators, consultants, study staff, and applicable Independent Ethics Committees National Competent Authorities or Institutional Review Boards. The contents of this document shall not be disclosed to others without written authorization from Sponsor.*

| Slides | Fixative | Evaluated organ | How do you regard the structural preservation of the tissue | How do you regard the preservation of the nuclei | How do you regard the preservation of the cytoplasm | How do you regard the diagnostic value of these preparations |
|---|---|---|---|---|---|---|
| 15 | PBF | Colon | Valid | Valid | Valid | Valid |
| 16 | GAF | Breast | Valid | Valid | Valid | Valid |
| 16 | PBF | Breast | Valid | Valid | Valid | Valid |
| 16 | GAF | Colon | Valid | Valid | Valid | Valid |
| 16 | PBF | Colon | Valid | Valid | Valid | Valid |
| 17 | GAF | Breast | Valid | Valid | Valid | Valid |
| 17 | PBF | Breast | Valid | Valid | Valid | Valid |
| 17 | GAF | Colon | Valid | Valid | Valid | Valid |
| 17 | PBF | Colon | Valid | Valid | Valid | Valid |
| 18 | GAF | Colon | Valid | Valid | Invalid | Valid |
| 18 | PBF | Colon | Valid | Valid | Valid | Valid |
| 19 | GAF | Breast | Valid | Valid | Valid | Valid |
| 19 | PBF | Breast | Valid | Valid | Valid | Valid |
| 19 | GAF | Colon | Valid | Valid | Invalid | Valid |
| 19 | PBF | Colon | Valid | Valid | Valid | Valid |
| 20 | GAF | Breast | Valid | Valid | Valid | Valid |
| 20 | PBF | Breast | Valid | Valid | Valid | Valid |
| 20 | GAF | Colon | Valid | Valid | Invalid | Valid |
| 20 | PBF | Colon | Valid | Valid | Valid | Valid |
| 21 | GAF | Breast | Valid | Valid | Valid | Valid |
| 21 | PBF | Breast | Valid | Valid | Valid | Valid |
| 21 | GAF | Colon | Valid | Valid | Invalid | Valid |
| 21 | PBF | Colon | Valid | Valid | Valid | Valid |
| 22 | GAF | Breast | Valid | Valid | Valid | Valid |
| 22 | PBF | Breast | Valid | Valid | Valid | Valid |

*This document is confidential and is to be distributed for review only to investigators, consultants, study staff, and applicable Independent Ethics Committees National Competent Authorities or Institutional Review Boards. The contents of this document shall not be disclosed to others without written authorization from Sponsor.*

| Slides | Fixative | Evaluated organ | How do you regard the structural preservation of the tissue | How do you regard the preservation of the nuclei | How do you regard the preservation of the cytoplasm | How do you regard the diagnostic value of these preparations |
|---|---|---|---|---|---|---|
| 22 | GAF | Colon | Valid | Valid | Invalid | Valid |
| 22 | PBF | Colon | Valid | Valid | Valid | Valid |
| 23 | GAF | Breast | Valid | Valid | Valid | Valid |
| 23 | PBF | Breast | Valid | Valid | Valid | Valid |
| 23 | GAF | Colon | Valid | Valid | Valid | Valid |
| 23 | PBF | Colon | Valid | Valid | Valid | Valid |
| 24 | GAF | Breast | Valid | Valid | Valid | Valid |
| 24 | PBF | Breast | Valid | Valid | Valid | Valid |
| 24 | GAF | Colon | Valid | Valid | Valid | Valid |
| 24 | PBF | Colon | Valid | Valid | Valid | Valid |
| 25 | GAF | Breast | Valid | Valid | Valid | Valid |
| 25 | PBF | Breast | Valid | Valid | Valid | Valid |
| 25 | GAF | Colon | Valid | Valid | Invalid | Valid |
| 25 | PBF | Colon | Valid | Valid | Valid | Valid |
| 26 | GAF | Breast | Valid | Valid | Valid | Valid |
| 26 | PBF | Breast | Valid | Valid | Valid | Valid |
| 26 | GAF | Colon | Valid | Valid | Valid | Valid |
| 26 | PBF | Colon | Valid | Valid | Valid | Valid |
| 27 | GAF | Breast | Valid | Valid | Valid | Valid |
| 27 | PBF | Breast | Valid | Valid | Valid | Valid |
| 27 | GAF | Colon | Valid | Valid | Valid | Valid |
| 27 | PBF | Colon | Valid | Valid | Valid | Valid |
| 28 | GAF | Breast | Valid | Valid | Valid | Valid |
| 28 | PBF | Breast | Valid | Valid | Valid | Valid |
| 29 | GAF | Breast | Valid | Valid | Valid | Valid |

*This document is confidential and is to be distributed for review only to investigators, consultants, study staff, and applicable Independent Ethics Committees National Competent Authorities or Institutional Review Boards. The contents of this document shall not be disclosed to others without written authorization from Sponsor.*

| Slides | Fixative | Evaluated organ | How do you regard the structural preservation of the tissue | How do you regard the preservation of the nuclei | How do you regard the preservation of the cytoplasm | How do you regard the diagnostic value of these preparations |
|---|---|---|---|---|---|---|
| 29 | PBF | Breast | Valid | Valid | Valid | Valid |
| 30 | GAF | Breast | Valid | Valid | Valid | Valid |
| 30 | PBF | Breast | Valid | Valid | Valid | Valid |
| 31 | GAF | Breast | Valid | Valid | Valid | Valid |
| 31 | PBF | Breast | Valid | Valid | Valid | Invalid |
| 33 | GAF | Breast | Valid | Valid | Valid | Valid |
| 33 | PBF | Breast | Valid | Valid | Valid | Valid |
| 34 | GAF | Breast | Valid | Valid | Valid | Valid |
| 34 | PBF | Breast | Valid | Valid | Valid | Valid |
| 35 | GAF | Breast | Valid | Valid | Valid | Valid |
| 35 | PBF | Breast | Valid | Valid | Valid | Valid |
| 36 | GAF | Breast | Valid | Valid | Valid | Valid |
| 36 | PBF | Breast | Valid | Valid | Valid | Valid |
| 37 | GAF | Breast | Valid | Valid | Valid | Valid |
| 37 | PBF | Breast | Valid | Valid | Valid | Valid |
| 38 | GAF | Breast | Valid | Valid | Valid | Valid |
| 38 | PBF | Breast | Valid | Valid | Valid | Valid |
| 40 | GAF | Breast | Valid | Valid | Valid | Valid |
| 40 | PBF | Breast | Valid | Valid | Valid | Valid |
| 41 | GAF | Breast | Valid | Valid | Valid | Valid |
| 41 | PBF | Breast | Valid | Valid | Valid | Valid |
| 42 | GAF | Breast | Valid | Valid | Valid | Valid |
| 42 | PBF | Breast | Valid | Valid | Valid | Valid |
| 43 | GAF | Breast | Valid | Valid | Valid | Valid |
| 43 | PBF | Breast | Valid | Valid | Valid | Valid |

*This document is confidential and is to be distributed for review only to investigators, consultants, study staff, and applicable Independent Ethics Committees National Competent Authorities or Institutional Review Boards. The contents of this document shall not be disclosed to others without written authorization from Sponsor.*

| Slides | Fixative | Evaluated organ | How do you regard the structural preservation of the tissue | How do you regard the preservation of the nuclei | How do you regard the preservation of the cytoplasm | How do you regard the diagnostic value of these preparations |
|---|---|---|---|---|---|---|
| 44 | GAF | Breast | Valid | Valid | Valid | Valid |
| 44 | PBF | Breast | Valid | Valid | Valid | Valid |
| 45 | GAF | Breast | Valid | Valid | Valid | Valid |
| 45 | PBF | Breast | Valid | Valid | Valid | Valid |
| 46 | GAF | Breast | Valid | Valid | Valid | Valid |
| 46 | PBF | Breast | Valid | Valid | Valid | Valid |
| 47 | GAF | Breast | Valid | Valid | Valid | Valid |
| 47 | PBF | Breast | Valid | Valid | Valid | Valid |
| 48 | GAF | Breast | Valid | Valid | Valid | Valid |
| 48 | PBF | Breast | Valid | Valid | Valid | Valid |
| 50 | GAF | Breast | Valid | Valid | Valid | Valid |
| 50 | PBF | Breast | Valid | Valid | Valid | Valid |
| 51 | GAF | Breast | Valid | Valid | Valid | Valid |
| 51 | PBF | Breast | Valid | Valid | Valid | Valid |
| 52 | GAF | Breast | Valid | Valid | Valid | Valid |
| 52 | PBF | Breast | Valid | Valid | Valid | Valid |

**Listing 3b**. Secondary Efficacy Endpoints

*This document is confidential and is to be distributed for review only to investigators, consultants, study staff, and applicable Independent Ethics Committees National Competent Authorities or Institutional Review Boards. The contents of this document shall not be disclosed to others without written authorization from Sponsor.*

| Slides | Fixative | Center | Evaluated organ | Do you consider that the preparations obtained on the same case with the two fixatives have the same performance? | Were you satisfied with the use of GAF fixative during the fixation procedure? | Local center pathologists Total score |
|---|---|---|---|---|---|---|
| 01 | GAF | Barcelona | Colon | No | 7.5 | 4 |
| 01 | PBF | Barcelona | Colon | . | . | 4 |
| 01 | GAF | Barcelona | Lung | Yes | 10 | 3 |
| 01 | PBF | Barcelona | Lung | . | . | 4 |
| 01 | GAF | Manchester | Endometrium | Yes | 10 | 4 |
| 01 | PBF | Manchester | Endometrium | . | . | 4 |
| 02 | GAF | Candiolo | Breast | Yes | 10 | 4 |
| 02 | PBF | Candiolo | Breast | . | . | 4 |
| 02 | GAF | Barcelona | Colon | Yes | 10 | 4 |
| 02 | PBF | Barcelona | Colon | . | . | 4 |
| 02 | GAF | Barcelona | Lung | Yes | 10 | 4 |
| 02 | PBF | Barcelona | Lung | . | . | 4 |
| 02 | GAF | Manchester | Endometrium | Yes | 10 | 4 |
| 02 | PBF | Manchester | Endometrium | . | . | 4 |
| 03 | GAF | Candiolo | Breast | Yes | 8.5 | 4 |
| 03 | PBF | Candiolo | Breast | . | . | 4 |
| 03 | GAF | Barcelona | Colon | Yes | 10 | 4 |
| 03 | PBF | Barcelona | Colon | . | . | 4 |
| 03 | GAF | Barcelona | Lung | Yes | 10 | 4 |
| 03 | PBF | Barcelona | Lung | . | . | 4 |
| 04 | GAF | Candiolo | Breast | Yes | 8 | 4 |
| 04 | PBF | Candiolo | Breast | . | . | 4 |

*This document is confidential and is to be distributed for review only to investigators, consultants, study staff, and applicable Independent Ethics Committees National Competent Authorities or Institutional Review Boards. The contents of this document shall not be disclosed to others without written authorization from Sponsor.*

| Slides | Fixative | Center | Evaluated organ | Do you consider that the preparations obtained on the same case with the two fixatives have the same performance? | Were you satisfied with the use of GAF fixative during the fixation procedure? | Local center pathologists Total score |
|---|---|---|---|---|---|---|
| 04 | GAF | Barcelona | Colon | Yes | 10 | 4 |
| 04 | PBF | Barcelona | Colon | . | . | 4 |
| 04 | GAF | Barcelona | Lung | Yes | 8 | 4 |
| 04 | PBF | Barcelona | Lung | . | . | 4 |
| 04 | GAF | Manchester | Endometrium | No | 6.3 | 4 |
| 04 | PBF | Manchester | Endometrium | . | . | 4 |
| 04 | GAF | Manchester | Prostate | Yes | 10 | 4 |
| 04 | PBF | Manchester | Prostate | . | . | 4 |
| 05 | GAF | Barcelona | Colon | Yes | 10 | 4 |
| 05 | PBF | Barcelona | Colon | . | . | 4 |
| 05 | GAF | Barcelona | Lung | Yes | 10 | 4 |
| 05 | PBF | Barcelona | Lung | . | . | 4 |
| 05 | GAF | Manchester | Endometrium | Yes | 10 | 4 |
| 05 | PBF | Manchester | Endometrium | . | . | 4 |
| 06 | GAF | Candiolo | Breast | Yes | 10 | 4 |
| 06 | PBF | Candiolo | Breast | . | . | 4 |
| 06 | GAF | Barcelona | Colon | Yes | 10 | 4 |
| 06 | PBF | Barcelona | Colon | . | . | 4 |
| 06 | GAF | Barcelona | Lung | Yes | 10 | 4 |
| 06 | PBF | Barcelona | Lung | . | . | 4 |
| 06 | GAF | Manchester | Prostate | No | 5.4 | 1 |
| 06 | PBF | Manchester | Prostate | . | . | 4 |

*This document is confidential and is to be distributed for review only to investigators, consultants, study staff, and applicable Independent Ethics Committees National Competent Authorities or Institutional Review Boards. The contents of this document shall not be disclosed to others without written authorization from Sponsor.*

| Slides | Fixative | Center | Evaluated organ | Do you consider that the preparations obtained on the same case with the two fixatives have the same performance? | Were you satisfied with the use of GAF fixative during the fixation procedure? | Local center pathologists Total score |
|---|---|---|---|---|---|---|
| 07 | GAF | Candiolo | Breast | No | 7 | 4 |
| 07 | PBF | Candiolo | Breast | . | . | 4 |
| 07 | GAF | Barcelona | Colon | Yes | 10 | 4 |
| 07 | PBF | Barcelona | Colon | . | . | 4 |
| 07 | GAF | Barcelona | Lung | Yes | 10 | 4 |
| 07 | PBF | Barcelona | Lung | . | . | 4 |
| 07 | GAF | Manchester | Endometrium | Yes | 10 | 3 |
| 07 | PBF | Manchester | Endometrium | . | . | 4 |
| 07 | GAF | Manchester | Prostate | No | 6.3 | 3 |
| 07 | PBF | Manchester | Prostate | . | . | 4 |
| 08 | GAF | Candiolo | Breast | Yes | 8.5 | 4 |
| 08 | PBF | Candiolo | Breast | . | . | 4 |
| 08 | GAF | Barcelona | Colon | Yes | 10 | 4 |
| 08 | PBF | Barcelona | Colon | . | . | 4 |
| 08 | GAF | Barcelona | Lung | Yes | 10 | 4 |
| 08 | PBF | Barcelona | Lung | . | . | 4 |
| 08 | GAF | Manchester | Endometrium | Yes | 10 | 4 |
| 08 | PBF | Manchester | Endometrium | . | . | 4 |
| 08 | GAF | Manchester | Prostate | No | 7.7 | 3 |
| 08 | PBF | Manchester | Prostate | . | . | 4 |
| 09 | GAF | Candiolo | Breast | No | 9 | 4 |
| 09 | PBF | Candiolo | Breast | . | . | 4 |

*This document is confidential and is to be distributed for review only to investigators, consultants, study staff, and applicable Independent Ethics Committees National Competent Authorities or Institutional Review Boards. The contents of this document shall not be disclosed to others without written authorization from Sponsor.*

| Slides | Fixative | Center | Evaluated organ | Do you consider that the preparations obtained on the same case with the two fixatives have the same performance? | Were you satisfied with the use of GAF fixative during the fixation procedure? | Local center pathologists Total score |
|---|---|---|---|---|---|---|
| 09 | GAF | Barcelona | Colon | No | 10 | 3 |
| 09 | PBF | Barcelona | Colon | . | . | 4 |
| 09 | GAF | Manchester | Endometrium | Yes | 10 | 4 |
| 09 | PBF | Manchester | Endometrium | . | . | 4 |
| 09 | GAF | Manchester | Prostate | No | 8.6 | 3 |
| 09 | PBF | Manchester | Prostate | . | . | 4 |
| 10 | GAF | Barcelona | Colon | Yes | 10 | 4 |
| 10 | PBF | Barcelona | Colon | . | . | 4 |
| 10 | GAF | Manchester | Endometrium | Yes | 10 | 4 |
| 10 | PBF | Manchester | Endometrium | . | . | 4 |
| 10 | GAF | Manchester | Prostate | No | 8.3 | 3 |
| 10 | PBF | Manchester | Prostate | . | . | 4 |
| 11 | GAF | Candiolo | Breast | Yes | 10 | 4 |
| 11 | PBF | Candiolo | Breast | . | . | 4 |
| 11 | GAF | Manchester | Endometrium | Yes | 5.5 | 3 |
| 11 | PBF | Manchester | Endometrium | . | . | 4 |
| 11 | GAF | Manchester | Prostate | No | 9.6 | 3 |
| 11 | PBF | Manchester | Prostate | . | . | 4 |
| 12 | GAF | Candiolo | Breast | Yes | 7 | 4 |
| 12 | PBF | Candiolo | Breast | . | . | 4 |
| 12 | GAF | Barcelona | Colon | Yes | 10 | 4 |
| 12 | PBF | Barcelona | Colon | . | . | 4 |

*This document is confidential and is to be distributed for review only to investigators, consultants, study staff, and applicable Independent Ethics Committees National Competent Authorities or Institutional Review Boards. The contents of this document shall not be disclosed to others without written authorization from Sponsor.*

| Slides | Fixative | Center | Evaluated organ | Do you consider that the preparations obtained on the same case with the two fixatives have the same performance? | Were you satisfied with the use of GAF fixative during the fixation procedure? | Local center pathologists Total score |
|---|---|---|---|---|---|---|
| 12 | GAF | Manchester | Endometrium | Yes | 8.4 | 3 |
| 12 | PBF | Manchester | Endometrium | . | . | 4 |
| 12 | GAF | Manchester | Prostate | No | 9.5 | 3 |
| 12 | PBF | Manchester | Prostate | . | . | 4 |
| 13 | GAF | Candiolo | Breast | Yes | 9 | 4 |
| 13 | PBF | Candiolo | Breast | . | . | 4 |
| 13 | GAF | Barcelona | Colon | Yes | 10 | 4 |
| 13 | PBF | Barcelona | Colon | . | . | 4 |
| 13 | GAF | Manchester | Endometrium | Yes | 10 | 4 |
| 13 | PBF | Manchester | Endometrium | . | . | 4 |
| 13 | GAF | Manchester | Prostate | No | 9.6 | 3 |
| 13 | PBF | Manchester | Prostate | . | . | 4 |
| 14 | GAF | Candiolo | Breast | Yes | 9 | 4 |
| 14 | PBF | Candiolo | Breast | . | . | 4 |
| 14 | GAF | Barcelona | Colon | Yes | 10 | 4 |
| 14 | PBF | Barcelona | Colon | . | . | 4 |
| 14 | GAF | Manchester | Prostate | No | 8.7 | 3 |
| 14 | PBF | Manchester | Prostate | . | . | 4 |
| 15 | GAF | Candiolo | Breast | Yes | 10 | 4 |
| 15 | PBF | Candiolo | Breast | . | . | 4 |
| 15 | GAF | Barcelona | Colon | Yes | 10 | 4 |
| 15 | PBF | Barcelona | Colon | . | . | 4 |

*This document is confidential and is to be distributed for review only to investigators, consultants, study staff, and applicable Independent Ethics Committees National Competent Authorities or Institutional Review Boards. The contents of this document shall not be disclosed to others without written authorization from Sponsor.*

| Slides | Fixative | Center | Evaluated organ | Do you consider that the preparations obtained on the same case with the two fixatives have the same performance? | Were you satisfied with the use of GAF fixative during the fixation procedure? | Local center pathologists Total score |
|---|---|---|---|---|---|---|
| 16 | GAF | Candiolo | Breast | Yes | 9 | 4 |
| 16 | PBF | Candiolo | Breast | . | . | 4 |
| 16 | GAF | Barcelona | Colon | Yes | 10 | 4 |
| 16 | PBF | Barcelona | Colon | . | . | 4 |
| 17 | GAF | Candiolo | Breast | Yes | 8.5 | 4 |
| 17 | PBF | Candiolo | Breast | . | . | 4 |
| 17 | GAF | Barcelona | Colon | No | 7 | 4 |
| 17 | PBF | Barcelona | Colon | . | . | 4 |
| 18 | GAF | Barcelona | Colon | No | 10 | 3 |
| 18 | PBF | Barcelona | Colon | . | . | 4 |
| 19 | GAF | Candiolo | Breast | Yes | 8.5 | 4 |
| 19 | PBF | Candiolo | Breast | . | . | 4 |
| 19 | GAF | Barcelona | Colon | No | 10 | 3 |
| 19 | PBF | Barcelona | Colon | . | . | 4 |
| 20 | GAF | Candiolo | Breast | Yes | 9 | 4 |
| 20 | PBF | Candiolo | Breast | . | . | 4 |
| 20 | GAF | Barcelona | Colon | Yes | 10 | 3 |
| 20 | PBF | Barcelona | Colon | . | . | 4 |
| 21 | GAF | Candiolo | Breast | Yes | 9.5 | 4 |
| 21 | PBF | Candiolo | Breast | . | . | 4 |
| 21 | GAF | Barcelona | Colon | Yes | 10 | 3 |
| 21 | PBF | Barcelona | Colon | . | . | 4 |

*This document is confidential and is to be distributed for review only to investigators, consultants, study staff, and applicable Independent Ethics Committees National Competent Authorities or Institutional Review Boards. The contents of this document shall not be disclosed to others without written authorization from Sponsor.*

| Slides | Fixative | Center | Evaluated organ | Do you consider that the preparations obtained on the same case with the two fixatives have the same performance? | Were you satisfied with the use of GAF fixative during the fixation procedure? | Local center pathologists Total score |
|---|---|---|---|---|---|---|
| 22 | GAF | Candiolo | Breast | Yes | 9 | 4 |
| 22 | PBF | Candiolo | Breast | . | . | 4 |
| 22 | GAF | Barcelona | Colon | No | 10 | 3 |
| 22 | PBF | Barcelona | Colon | . | . | 4 |
| 23 | GAF | Candiolo | Breast | Yes | 10 | 4 |
| 23 | PBF | Candiolo | Breast | . | . | 4 |
| 23 | GAF | Barcelona | Colon | Yes | 10 | 4 |
| 23 | PBF | Barcelona | Colon | . | . | 4 |
| 24 | GAF | Candiolo | Breast | Yes | 9 | 4 |
| 24 | PBF | Candiolo | Breast | . | . | 4 |
| 24 | GAF | Barcelona | Colon | Yes | 10 | 4 |
| 24 | PBF | Barcelona | Colon | . | . | 4 |
| 25 | GAF | Candiolo | Breast | Yes | 9 | 4 |
| 25 | PBF | Candiolo | Breast | . | . | 4 |
| 25 | GAF | Barcelona | Colon | Yes | 10 | 3 |
| 25 | PBF | Barcelona | Colon | . | . | 4 |
| 26 | GAF | Candiolo | Breast | Yes | 10 | 4 |
| 26 | PBF | Candiolo | Breast | . | . | 4 |
| 26 | GAF | Barcelona | Colon | Yes | 10 | 4 |
| 26 | PBF | Barcelona | Colon | . | . | 4 |
| 27 | GAF | Candiolo | Breast | Yes | 9 | 4 |
| 27 | PBF | Candiolo | Breast | . | . | 4 |

*This document is confidential and is to be distributed for review only to investigators, consultants, study staff, and applicable Independent Ethics Committees National Competent Authorities or Institutional Review Boards. The contents of this document shall not be disclosed to others without written authorization from Sponsor.*

| Slides | Fixative | Center | Evaluated organ | Do you consider that the preparations obtained on the same case with the two fixatives have the same performance? | Were you satisfied with the use of GAF fixative during the fixation procedure? | Local center pathologists Total score |
|---|---|---|---|---|---|---|
| 27 | GAF | Barcelona | Colon | Yes | 10 | 4 |
| 27 | PBF | Barcelona | Colon | . | . | 4 |
| 28 | GAF | Candiolo | Breast | Yes | 9 | 4 |
| 28 | PBF | Candiolo | Breast | . | . | 4 |
| 29 | GAF | Candiolo | Breast | Yes | 8.5 | 4 |
| 29 | PBF | Candiolo | Breast | . | . | 4 |
| 30 | GAF | Candiolo | Breast | Yes | 9.5 | 4 |
| 30 | PBF | Candiolo | Breast | . | . | 4 |
| 31 | GAF | Candiolo | Breast | Yes | 8.6 | 4 |
| 31 | PBF | Candiolo | Breast | . | . | 3 |
| 33 | GAF | Candiolo | Breast | Yes | 9 | 4 |
| 33 | PBF | Candiolo | Breast | . | . | 4 |
| 34 | GAF | Candiolo | Breast | Yes | 9.5 | 4 |
| 34 | PBF | Candiolo | Breast | . | . | 4 |
| 35 | GAF | Candiolo | Breast | Yes | 8.5 | 4 |
| 35 | PBF | Candiolo | Breast | . | . | 4 |
| 36 | GAF | Candiolo | Breast | Yes | 9.1 | 4 |
| 36 | PBF | Candiolo | Breast | . | . | 4 |
| 37 | GAF | Candiolo | Breast | Yes | 9 | 4 |
| 37 | PBF | Candiolo | Breast | . | . | 4 |
| 38 | GAF | Candiolo | Breast | Yes | 8.1 | 4 |
| 38 | PBF | Candiolo | Breast | . | . | 4 |

*This document is confidential and is to be distributed for review only to investigators, consultants, study staff, and applicable Independent Ethics Committees National Competent Authorities or Institutional Review Boards. The contents of this document shall not be disclosed to others without written authorization from Sponsor.*

| Slides | Fixative | Center | Evaluated organ | Do you consider that the preparations obtained on the same case with the two fixatives have the same performance? | Were you satisfied with the use of GAF fixative during the fixation procedure? | Local center pathologists Total score |
|---|---|---|---|---|---|---|
| 40 | GAF | Candiolo | Breast | Yes | 9.5 | 4 |
| 40 | PBF | Candiolo | Breast | . | . | 4 |
| 41 | GAF | Candiolo | Breast | Yes | 8.5 | 4 |
| 41 | PBF | Candiolo | Breast | . | . | 4 |
| 42 | GAF | Candiolo | Breast | Yes | 7.6 | 4 |
| 42 | PBF | Candiolo | Breast | . | . | 4 |
| 43 | GAF | Candiolo | Breast | Yes | 9 | 4 |
| 43 | PBF | Candiolo | Breast | . | . | 4 |
| 44 | GAF | Candiolo | Breast | Yes | 9 | 4 |
| 44 | PBF | Candiolo | Breast | . | . | 4 |
| 45 | GAF | Candiolo | Breast | Yes | 9 | 4 |
| 45 | PBF | Candiolo | Breast | . | . | 4 |
| 46 | GAF | Candiolo | Breast | Yes | 8 | 4 |
| 46 | PBF | Candiolo | Breast | . | . | 4 |
| 47 | GAF | Candiolo | Breast | Yes | 9 | 4 |
| 47 | PBF | Candiolo | Breast | . | . | 4 |
| 48 | GAF | Candiolo | Breast | Yes | 8.8 | 4 |
| 48 | PBF | Candiolo | Breast | . | . | 4 |
| 50 | GAF | Candiolo | Breast | Yes | 10 | 4 |
| 50 | PBF | Candiolo | Breast | . | . | 4 |
| 51 | GAF | Candiolo | Breast | Yes | 10 | 4 |
| 51 | PBF | Candiolo | Breast | . | . | 4 |

*This document is confidential and is to be distributed for review only to investigators, consultants, study staff, and applicable Independent Ethics Committees National Competent Authorities or Institutional Review Boards. The contents of this document shall not be disclosed to others without written authorization from Sponsor.*

| Slides | Fixative | Center | Evaluated organ | Do you consider that the preparations obtained on the same case with the two fixatives have the same performance? | Were you satisfied with the use of GAF fixative during the fixation procedure? | Local center pathologists Total score |
|---|---|---|---|---|---|---|
| 52 | GAF | Candiolo | Breast | Yes | 10 | 4 |
| 52 | PBF | Candiolo | Breast | . | . | 4 |

*This document is confidential and is to be distributed for review only to investigators, consultants, study staff, and applicable Independent Ethics Committees National Competent Authorities or Institutional Review Boards. The contents of this document shall not be disclosed to others without written authorization from Sponsor.*

### 8 LIST OF ABBREVATIONS AND DEFINITIONS

| | |
|---|---|
| AE | Adverse Event |
| ADE | Adverse Device Effect |
| CA/RA | Competent Authority / Regulatory Authority |
| CI | Confidence Interval |
| CPSP | Clinical Performance Study Protocol |
| CRA | Clinical Research Associate |
| CRF | Case Report Form |
| CSR | Clinical Study Report |
| EC | Ethics Committee |
| GCP | Good Clinical Practice |
| ICH | International Conference on Harmonization |
| IRB | Institutional Review Board |
| ISF | Investigator's Site File |
| ITT | Intention-To-Treat (population) |
| PI | Principal Investigator |
| PP | Per Protocol (population) |
| PVC | Polyvinyl Chloride |
| SADE | Serious Adverse Device Effect |
| SAE | Serious Adverse Event |
| SAP | Statistical Analysis Plan |
| SCR | Screening Number |
| SD | Standard Deviation |
| SOP | Standard Operating Procedure |
| UADE | Unanticipated Adverse Device Effect |

*This document is confidential and is to be distributed for review only to investigators, consultants, study staff, and applicable Independent Ethics Committees National Competent Authorities or Institutional Review Boards. The contents of this document shall not be disclosed to others without written authorization from Sponsor.*

### 9 ETHICS

The study was conducted in compliance with the independent Ethics Committee/Institutional Review Board (EC/IRB)'s recommendation, informed consent regulations (written Informed Consent was obtained in response to a fully written and verbal explanation of the nature of the study prior to start any procedure scheduled for the study), Declaration of Helsinki, Good Clinical Practice guidelines, local laws, ISO 20916:2019 and relative Study Protocol. In addition, the study adhered to all applicable local and international laws and regulation.

| | Site 1 | Site 2 | Site 3 |
|---|---|---|---|
| **Ethics committee / IRB** | 22.10.2019/NA | 15.10.2019/NA | 16.01.2020/NA |
| **Date final protocol approved by EC** | 19.09.2019_V1 | 19.09.2019_V1 | 19.09.2019_V1 |

### 10 INVESTIGATORS AND ADMINISTRATIVE STRUCTURE OF STUDY

#### 10.1 PRINCIPAL INVESTIGATOR(S)

As per study design, this is a multicenter study, the centers are below reported including the Coordinating Investigator (*):

Three European Institutions are involved for the sampling:
Istituto per la Ricerca e cura del Cancro (Institute for Cancer Research and Cure, IRCCS of Candiolo (Torino, Italy)). Strada Provinciale 142 km 39,5 - 10060 Candiolo (TO).
PI: Prof. Anna Sapino (*) (Scientific Director of the Institute, Head of the Service of Pathological Anatomy and Histology) - Tel. +39-011-9933201-3211.

Hospital Universitari Vall d'Hebron; Vall d'Hebron Barcelona Hospital Campus Passeig de la Vall d'Hebron, 119-129 - 08035 Barcelona (Spain)
PI: Prof. Santiago Ramon y Cajal (Head of Pathology Service) - Tel. +34 934893000 (Ext. 6934).

The Christie NHS Foundation Trust Wilmslow Road, Manchester, M20 4BX. United Kingdom.
PI: Dr. Pedro Soares de Oliveira consultant in histopathology. Dept. Of pathology. - tel. +44-161-4463275

#### 10.2 EXTERNAL ORGANIZATIONS INVOLVED (CRO, LABORSTORIES, CONSULTANTS)

1MED SA (CRO)
Address: Via Campagna, 13
6982 Agno – CH

1MED supported the Sponsor with the following activities:

- Study Planning, preparation, and project management.

*This document is confidential and is to be distributed for review only to investigators, consultants, study staff, and applicable Independent Ethics Committees National Competent Authorities or Institutional Review Boards. The contents of this document shall not be disclosed to others without written authorization from Sponsor.*

- Study Initiation and Submission.
- Monitoring activities.
- Data management.
- Biostatistics.
- Medical Writing.

Reading of the slides will be performed by the Central Pathology Reviewer: Prof. Ales Ryska Charles University Hradec Kralove, Czech Republic.

#### 10.3 SPONSOR INFORMATION

ADDAX Biosciences Srl,
Strada Mongreno 247,
10132 Torino, Italy

*This document is confidential and is to be distributed for review only to investigators, consultants, study staff, and applicable Independent Ethics Committees National Competent Authorities or Institutional Review Boards. The contents of this document shall not be disclosed to others without written authorization from Sponsor.*

*This document is confidential and is to be distributed for review only to investigators, consultants, study staff, and applicable Independent Ethics Committees National Competent Authorities or Institutional Review Boards. The contents of this document shall not be disclosed to others without written authorization from Sponsor.*

13. TOXNET–Toxicology Data Network. GLYOXAL (CASRN: 107-22-2). https://toxnet.nlm.nih.gov/cgi- bin/sis/search/a?dbs+hsdb:@term+@DOCNO+497
14. World Health Organization/International Programme on Chemical Safety. Concise International Chemi- cal Assessment Document No. 57 Glyoxal. 2004.
15. Sabatini DD, Bensch K, Barrnett RJ. Cytochemistry and electron microscopy. The preservation of cellu- lar ultrastructure and enzymatic activity by aldehyde fixation. J Cell Biol. 1963; 17:19–58.
16. Hopwood D. The elution patterns of formaldehyde, glutaraldehyde, glyoxal and alpha-hydroxyadipalde- hyde from sephadex G-10 and their significance for tissue fixation. Histochemie. 1969; 20(2):127–32.
17. Dapson RW. Glyoxal fixation: how it works and why it only occasionally needs antigen retrieval. Biotech Histochem. 2007; 82(3):161–6.
18. Buesa RJ. Histology without formalin? Ann Diagn Pathol. 2008; 12(6):387–96.
19. Marcon N, Bressenot A, et al. Le glyoxal: un possible substitut polyvalent du formalde´ hyde en anatomie pathologique? [Glyoxal: a possible polyva- lent substitute for formaldehyde in pathology?]. Ann Pathol. 2009; 29(6):460–7.
20. Umlas J, Tulecke M. The effects of glyoxal fixation on the histological evaluation of breast specimens. Hum Pathol. 2004; 35(9):1058–62.
21. Tubbs RR, Hsi ED, Hicks D, Goldblum J. Molecular pathology testing of tissues fixed in prefer solution. Am J Surg Pathol. 2004; 28(3):417–9.
22. Willmore-Payne C, Metzger K, Layfield LJ. Effects of fixative and fixation protocols on assessment of Her-2/neu oncogene amplification status by fluorescence in situ hybridization. Appl Immunohistochem Mol Morphol. 2007; 15(1):84–7.
23. Lassalle S, Hofman V, Marius I, Gavric-Tanga V, Brest P, Havet K, et al. Assessment of morphology, antigenicity, and nucleic acid integrity for diagnostic thyroid pathology using formalin substitute fixatives. Thyroid. 2009; 19(11):1239–48.
24. Gillespie JW, Best CJ, Bichsel VE, Cole KA, Greenhut SF, Hewitt SM, et al. Evaluation of non-formalin tissue fixation for molecular profiling studies. Am J Pathol. 2002; 160(2):449–57.
25. Foss RD, Guha-Thakurta N, Conran RM, Gutman P. Effects of fixative and fixation time

*This document is confidential and is to be distributed for review only to investigators, consultants, study staff, and applicable Independent Ethics Committees National Competent Authorities or Institutional Review Boards. The contents of this document shall not be disclosed to others without written authorization from Sponsor.*

on the extrac- tion and polymerase chain reaction amplification of RNA from paraffin-embedded tissue. Comparison of two housekeeping gene mRNA controls. Diagn Mol Pathol. 1994; 3(3):148–55.

26. Zhang Z, Zhao D, Xu B. Analysis of glyoxal and related substances by means of high-performance liq- uid chromatography with refractive index detection. J Chromatogr Sci. 2013; 51(10):893–8.
27. Conroy R. Sample size: a rough guide.2006. http://www.beaumontethics.ie/docs/application/samplesizecalculation.pdf.Accessed November 16, 2013.
28. SAS. Institute Inc., Cary, North Carolina, United States of America, Version 9.4.

*This document is confidential and is to be distributed for review only to investigators, consultants, study staff, and applicable Independent Ethics Committees National Competent Authorities or Institutional Review Boards. The contents of this document shall not be disclosed to others without written authorization from Sponsor.*

### 12 ANNEXES

#### 12.1 CPSP (INCL. AMENDMENTS)

#### 12.2 IFU

#### 12.3 PRINICIPAL INVESTIGATOR(S)

#### 12.4 EXTERNAL ORGANIZATIONS

#### 12.5 AUDIT CERTIFICATE

No audit performed during the CPS

*This document is confidential and is to be distributed for review only to investigators, consultants, study staff, and applicable Independent Ethics Committees National Competent Authorities or Institutional Review Boards. The contents of this document shall not be disclosed to others without written authorization from Sponsor.*
